# Prediction models of bronchopulmonary dysplasia: a systematic review and meta-analysis with validation

**DOI:** 10.1101/2022.07.19.22277664

**Authors:** T’ng Chang Kwok, Natalie Batey, Ka Ling Luu, Andrew Prayle, Don Sharkey

## Abstract

**Introduction:** Neonatal bronchopulmonary dysplasia (BPD) is associated with lifelong respiratory and neurological sequalae. Prediction models could identify infants at greatest risk of BPD and allow targeted preventative strategies. We performed a systematic review and meta-analysis with external validation of identified models.

**Methods:** Studies using predictors available before day 14 of life to predict BPD in very preterm infants were included. Two reviewers assessed 7,628 studies for eligibility. Meta-analysis of externally validated models was followed by validation using 62,864 very preterm infants in England and Wales.

**Results:** 64 studies using 53 prediction models were included totalling 274,407 infants (range 32–156,587/study). 35 (55%) studies predated 2010; 39 (61%) were single-centre studies. 46 (87%) models were developed for the first week of life. Overall, 97% of studies had a high risk of bias, especially in the analysis domain. Internal (25%) and external (30%) validation were performed infrequently in the 44 model derivation studies. Following meta-analysis of 22 BPD and 11 BPD/death composite models, Laughon’s day one model was the most promising in predicting BPD and death with a fair C-statistic of 0.76 (95% CI 0.70–0.81) and good calibration. Six models were externally validated in our cohort with a C-statistic between 0.70 to 0.90 but with poor calibration.

**Conclusion:** Few BPD prediction models were developed with contemporary populations, underwent external validation, or had calibration and impact analyses. To reduce the adverse impact of BPD, we need contemporary, validated, and dynamic prediction models to allow targeted preventative strategies.

**What is the key question?:** This review aims to provide a comprehensive assessment of all BPD prediction models developed to address the clinical uncertainty of which predictive model is sufficiently valid and generalisable for use in clinical practice and research.

**What is the bottom line?:** Published BPD prediction models are mostly outdated, single centre and lack external validation.

**Why read on?:** Laughon’s 2011 model is the most promising but more robust models, using contemporary data with external validation are needed to support better treatments.

## Introduction

Bronchopulmonary dysplasia (BPD), one of the most common and complex neonatal conditions^1^, continues to increase and affects approximately 28,000 and 18,000 babies annually in Europe^2^ and the US^3^ respectively. Preterm infants with BPD have significant long-term respiratory and neurodevelopmental complications into adulthood^4^, including abnormal lung function^5^ and poor school performance^4^.

There are a myriad of trials with at least 24 Cochrane reviews looking at BPD preventative interventions, including postnatal corticosteroids. However, their benefit in preventing BPD may not outweigh the significant side effects, including gastrointestinal perforation and neurodevelopmental impairment^6 7^. This demonstrates the complexity in BPD management in balancing the risk of significant long-term morbidity from BPD with that of exposure to potentially harmful treatments^8^.

BPD prediction models aim to provide a personalised risk approach in identifying high-risk very preterm infants for timely preventative treatments. Despite numerous models being developed, none are used routinely in clinical practice. This review aims to provide a comprehensive assessment of all BPD prediction models developed to address the clinical uncertainty of which predictive model is sufficiently valid and generalisable for clinical and research use. Secondly, we will validate eligible models in a large national contemporaneous cohort of very preterm infants.

## Material and Methods

### Systematic Review

There was no deviation from the protocol published in PROSPERO^9^. Standard Cochrane Neonatal and Prognosis Methods Group methodology were used.

### Inclusion criteria

Cohort, case-control, and randomised controlled trials used in developing or validating the prediction models were included. Very preterm infants born before 32 weeks of gestational age (GA) and less than two weeks old at the time of BPD prediction were included. This ensures the clinical applicability and timeliness of the prediction models to support clinical decision making on preventative treatments. Studies that used non-universally accessible predictors such as pulmonary function tests, ultrasonography and biomarkers were excluded. BPD was defined as a respiratory support requirement at either 28 days of age or 36 weeks of corrected gestational age (CGA)^10^. The composite outcome of BPD and death before discharge was included as a secondary outcome.

### Search methods

Standard Cochrane Neonatal^11^ and prognostic study search filters^12^ were used. “Bronchopulmonary dysplasia OR BPD OR chronic lung disease OR CLD” search terms were used to search the CENTRAL, Ovid MEDLINE, CINAHL, EMBASE and Scopus databases until 13/08/2021 (**Appendix 1**).

### Data collection

Two reviewers (TK, NB or KL) independently screened the title and abstract as well as full-text reports for inclusion before independently extracting data and assessing the risk of bias using the PROBAST tool^13^ (**Appendix 2**). These were done using a web-based tool CADIMA^14^. Any disagreement was resolved by discussion.

### Prediction model performance measure

Discrimination (C-statistics), calibration (Observed:Expected ratio (O:E ratio)) and classification (net benefit analysis) measures were extracted alongside their uncertainties.

### Missing data

Study authors were contacted to obtain any missing data. Failing that, missing performance measure were approximated using the methodology proposed by Debray et al.^15^ and R statistical package “metamisc”^16^.

### Meta-analysis

Meta-analysis of the performance measures, using the random-effects approach and R statistical package “metafor”^17^, was performed for externally validated models. Sensitivity analysis was performed by excluding studies with an overall high risk of bias. We pre-specified that we would assess the source of heterogeneity^15^ and reporting deficiencies^18^ if more than ten studies were included.

### Conclusions

The adapted **<u>G</u>**rades of **<u>R</u>**ecommendation, **<u>A</u>**ssessment, **<u>D</u>**evelopment and **<u>E</u>**valuation (GRADE) framework^19^ was used to assess the certainty of the evidence.

## External Validation of Eligible Models

### Study design

A population-based retrospective cohort study from the UK National Neonatal Research Database (NNRD)^20^ was used to externally validate BPD prediction models identified in the systematic review. We included all very preterm infants admitted to 185 neonatal units in England and Wales from January 2010 to December 2017. This encompasses over 90% of English neonatal units in 2010, with full coverage in England and Wales in 2012 and 2014 respectively. Infants with birthweight z score below -4 or above 4 were excluded as they were likely erroneous entries. Further details of the data items extracted are found within the National Neonatal Dataset[29] and **Appendix 3**. Ethical approval was granted by the Sheffield Research Ethics Committee (REC reference 19/YH/0115).

### Statistical analysis

Data extraction and statistical analysis were done using STATA/SE version 16 (StataCorp) and R version 4 (R Core Team). Summary statistics (median, interquartile range (IQR) and percentages) were used to describe the data. Missing data was imputed five times using Multivariate Imputation by Chained Equations^21^. Model performances were assessed in three domains: discrimination (C-statistics), calibration (calibration plot and O:E ratio) and utility measure (decision curve analysis).

## Results

### Systematic review

#### Literature search

Of the 7,628 potentially eligible studies identified, 194 full-text articles were screened with 122 articles excluded as studies identified risk factors rather than developing prediction models (48%), predictors available after two weeks of age (24%), infants above 32 weeks GA at birth (17%), non-universally accessible predictors (10%) or wrong outcome measure reported (2%). Data were extracted from the 72 full-text articles (**Appendix 4**), encompassing 64 studies and 53 BPD prediction models (**Figure 1**).

**Figure 1:**
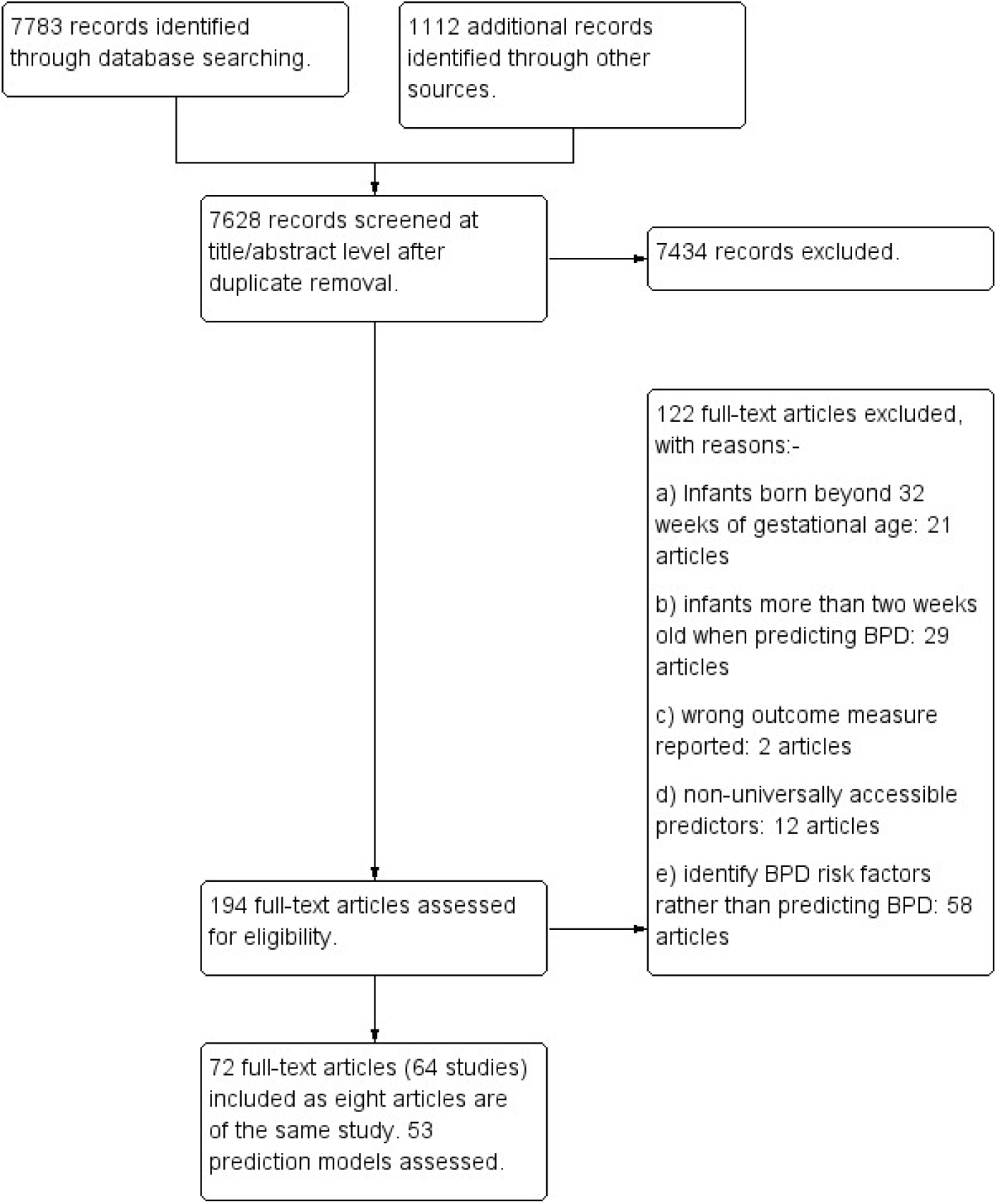
Flow diagram of literature search and included studies.

#### Description of included studies

Of the 64 included studies, 31 were BPD prediction model development studies, 20 were validation studies, and 13 were development and external validation studies. Fifty-five of the studies were cohort studies; five were randomised controlled trials; two used a combination of randomised control trials and cohort studies with one case-control study and another with unreported study design. Twenty-six studies were performed in North America, fourteen in Europe, thirteen in Asia, five in South America and Australia/New Zealand each and one study was carried out worldwide. Twenty studies developed and validated BPD prediction models based on infants born before 2000, with a further 15 studies using infants born between 2000 to 2010. The 64 included studies recruited 274,407 (range 32 and 156,587) infants, with the majority (50 studies) recruited less than 1,000 infants. 39 (61%) studies were conducted in single centre. Forty-seven studies used BPD as their outcome, while 14 studies used a BPD/death composite outcome, with three further studies reporting both BPD and BPD/death composite outcomes. Thirty-one studies defined BPD at 36 weeks CGA, while 22 studies used the timepoint of 28 days old. Six studies defined BPD using both timepoints. Five studies did not report how BPD was defined (**Table 1**).

**Table 1.** Characteristics of 64 included studies. NR = Not reported. D = Derivation. V = Validation. RCT = Randomised controlled trials. Sev= Severe. * Data obtained from the original study protocol (Cools et al 2010^22^).

| Study | Country / Region | Number of centres | Data collection period | Study design | Number of infants | Number of BPD | Number of deaths | Gestation | Birthweight (grams) | Outcome |
| --- | --- | --- | --- | --- | --- | --- | --- | --- | --- | --- |
| **Development only** |  |  |  |  |  |  |  |  |  |  |
| Cohen 1983 | USA | 1 | 1978 – 1981 | Cohort | 69 | 42 | NR | NR | NR | BPD 28d |
| Palta 1990 | USA | 5 | NR | Cohort | 42 | 36 | NR | NR | 1042 ± 267 | BPD 30d |
| Parker 1992 | USA | 1 | 1976 – 1985 (D) 1986 – 1990 (V) | Cohort | 1500 (D) 875 (V) | 252 (D) 288 (V) | 328 (D) 139 (V) | 29.6 ± 2.6 (D) 28.7 ± 2.9 (V) | 1113 ± 261 (D) 1066 ± 278 (V) | BPD 28d & Death/BPD28d |
| Corcoran 1993 | UK | 1 | 1980 – 1990 | Cohort | 412 | 140 | 115 | 29.7 ± 2.8 | 1345 ± 445 | BPD 28d |
| Gortner 1996 | Germany | 1 | 1985 – 1992 | Case – control | 152 | 76 | 48 | 29.2 ± 2.0 | 1139 ± 249 | BPD 28d |
| Ryan 1996 | UK | 1 | 1991 – 1992 (D) 1993 (V) | Cohort | 204 (D) 47 (V) | BPD 28d 85 (D) NR (V)<br>BPD 36w 51 (D) NR (V) | 7 (D) NR (V) | 27.5 ± 1.5 (D) NR (V) | 1283 ± 327 (D) NR (V) | BPD 28d & BPD 36w |
| Groves 2004 | NZ | 1 | 1998 – 2000 | Cohort | 290 | 60 | 54 | 27.9 ± 8.4 | 863 ± 626 | Death/BPD 36w |
| Cunha 2005 | Brazil | 1 | 2000 – 2002 | Cohort | 86 | 45 | NR | 29.0 ± 2.3 | 1029 ± 222 | BPD 28d |
| Choi 2006 | Korea | 1 | 2000 – 2005 | Cohort | 81 | 48 | NR | 28.1 ± 1.7 | 1051 ± 233 | BPD 28d |
| Henderson-Smart 2006 | Australia / NZ | 25 | 1998 – 1999 (D) 2000 – 2001 (V) | Cohort | 5599 (D) 5854 (V) | 1235 (D) 1475 (V) | NR | 28.7 ± 2.2 (D) 28.7 ± 2.2 (V) | 1233 ± 404 (D) 1235 ± 408 (V) | BPD 36w |
| Ambalavanan 2008 | USA | 16 | 2001 – 2003 | RCT | 420 | 151 | 202 | 26.0 ± 2.0 | 839 ± 262 | Death/BPD 36w |
| Gottipati 2012 | USA | 1 | 2002 – 2007 | Cohort | 417 | NR | NR | NR | NR | BPD (NR) |
| Roth-Kleiner 2012 | Switzerland | 1 | 1998 – 2007 | Cohort | 936 | NR | NR | NR | NR | BPD (NR) |
| Chock 2014 | USA | 1 | 2006 – 2010 | Cohort | 187 | 73 | 12 | 27.6 ± 2.0 | 1005 ± 260 | Death/BPD 36w |
| Yang 2014 | Korea | 1 | 2003 – 2010 | Cohort | 261 | 66 | 0 | 30.6 ± 2.4 | 1549 ± 487 | BPD 28d |
| Ochab 2016 | Poland | 1 | NR | NR | 109 | 46 | NR | NR | NR | BPD 28d |
| Wai 2016 | USA | 25 | 2010 – 2013 | RCT | 495 | 283 | 53 | 25.2 ± 1.2 | 700 ± 165 | Death/BPD 36w |
| Kim 2017 | Korea | 1 | 2008 – 2014 | Cohort | 304 | 110 | 13 | 28.3 ± 2.3 | 1032 ± 276 | Death/BPD 36w |
| Beltempo 2018 | Canada | 30 | 2010 – 2015 | Cohort | 9240 | 2959 | 1277 | 26.7 | 925 ± 251 | BPD 36w |
| Boghossian 2018 | USA / Puerto Rico | 852 | 2006 – 2014 | Cohort | 156587 | NR | NR | NR | NR | BPD 36w |
| Hunt 2018 | UK | 1 | 2012 – 2017 | Cohort | 432 | 228 | 7 | NR | NR | BPD 28d |
| Sullivan 2018 | USA | 2 | 2009 – 2015 | Cohort | 778 | 186 | 48 | 28.0 ± 2.8 | 1029 ± 298 | BPD 36w |
| Fairchild 2019 | USA | 1 | 2009 – 2014 | Cohort | 502 | 172 | 15 | 27.3 ± 3.0 | 1023 ± 335 | BPD 36w |
| Sun 2019 | China | 1 | 2015 – 2018 | Cohort | 296 | 144 | 0 | 29.9 ± 1.6 | 1417 ± 328 | BPD 28d |
| Valenzuela–Stutman 2019 | South America | 15 | 2001 – 2015 | Cohort | 16407 | 2580 | 3938 | 29 ± 2.9 | 1099 ± 275 | BPD 36w & Death/BPD 36w |
| Dylag 2020 | USA | 6 | 2011 – 2014 | Cohort | 704 | BPD 28d<br>414<br>BPD 36w<br>276 | 0 | 26.7 ± 1.4 | 922 ± 229 | BPD 28d |
| Shah 2020 | USA | 1 | 2006 – 2016 | Cohort | 730 | 343 | 139 | 27 ± 2 | 867 ± 198 | Death/BPD 36w |
| Sharma 2020 | USA | 1 | 2011 – 2017 | Cohort | 263 | 155 | 16 | 25.2 ± 1.4 | 805 ± 195 | BPD 36w |
| Vaid 2020 | USA | 1 | 2005 – 2018 | Cohort | 1832 | NR | NR | NR | NR | BPD (NR) |
| Shim 2021 | Korea | 66 | 2013 – 2016<br>(D) 2017 –<br>2017 (V) | Cohort | 4600 (D)<br>1740 (V) | BPD 28d<br>2583 (D)<br>1003 (V)<br>BPD 36w<br>1370 (D)<br>463 (V) | 1053 (D)<br>280 (V) | 28.7 ± 2.6 (D)<br>28.8 ± 2.6 (V) | 1119 ± 264 (D)<br>1127 ± 260 (V) | BPD 28d & BPD<br>36w |
| Ushida 2021 | Japan | 200 | 2006 – 2015 | Cohort | 31157 | 7504 | 1958 | 27.8 ± 2.5 | 973 ± 299 | BPD 36w |
| **<u>Development and validation</u>** |  |  |  |  |  |  |  |  |  |  |
| Ryan 1994 | UK | 2 | 1988 – 1989 | Cohort | 166 (D)<br>133 (V) | 47 (D)<br>59 (V) | NR | 29 ± 2.6 (D)<br>30 ± 3.1 (V) | 1043 ± 189 (D)<br>1056 ± 177 (V) | BPD 28d |
| Rozycki 1996 | USA | 1 | 1987 – 1989<br>(D) 1990 –<br>1991 (V) | Cohort | 14d model<br>116 (D) 61<br>(V)<br>8h model<br>698 (D) | 14d model<br>38 (D) 34 (V)<br>8h model<br>44 (D) | NR | 14d model<br>26.7 ± 2.0 (D)<br>NR(V)<br>8h model<br>29.7 ± 2.2 (D) | 14d model<br>911 ± 227 (D)<br>NR(V)<br>8h model<br>1352 ± 478 (D) | BPD 28d |
| Romagnoli 1998 | Italy | 1 | 1989 – 1991<br>(D) 1993 –<br>1996 (V) | Cohort | 50 (D)<br>149 (V) | 28 (D)<br>82 (V) | NR | 28.4 ± 2.1 (D)<br>28.7 ± 2.4 (V) | 893 ± 206 (D)<br>931 ± 208 (V) | BPD 28d |
| Yoder 1999 | USA | 1 (D)<br>3 (V) | 1990 – 1992<br>(D) 1993 –<br>1995 (V) | Cohort | 48 (D)<br>110 (V) | 15 (D)<br>33 (V) | NR | 27.0 ± 2.0 (D)<br>26.5 ± 2.1 (V) | 897 ± 243 (D)<br>905 ± 222 (V) | Death/BPD 36w |
| Chien 2002 | Canada | 17 | 1996 – 1997 | Cohort | 4226 | NR | NR | 29.0 ± 2.0 | 1390 ± 457 | BPD 36w |
| Kim 2005 | Korea | 1 | 1997 – 1999<br>(D) 2000 –<br>2001 (V) | Cohort | 197 (D)<br>107 (V) | 30 (D)<br>9 (V) | 34 (D)<br>11 (V) | 28.2 ± 1.9 (D)<br>28.5 ± 1.9 (V) | 1043 ± 263 (D)<br>1095 ± 270 (V) | BPD 36w |
| Bhering 2007 | Brazil | 1 | 1998 – 2003<br>(D) 2003 –<br>2005 (V) | Cohort | 247 (D)<br>61 (V) | 68 (D)<br>NR (V) | 5 (D)<br>NR (V) | 29.1 ± 2.4 (D)<br>NR (V) | 1083 ± 237 (D)<br>NR (V) | BPD 28d |
| May 2007 | UK | 1 | 1995 – 1998<br>(D) 2004 –<br>2005 (V) | RCT<br>(D)<br>Cohort<br>(V) | 136 (D)<br>75 (V) | BPD 28d<br>82 (D) 32 (V)<br>BPD 36w<br>38 (D) 22 (V) | NR | 27.7 ± 2.0 (D)<br>29.3 ± 2.6 (V) | 1017 ± 246 (D)<br>1245 ± 424 (V) | BPD 28d & BPD<br>36w |
| Laughon 2011 | USA | 17 | 2000 – 2004 | RCT | 3629 (D)<br>1777 (V) | 1943 (D)<br>1215 (V) | 468 (D)<br>210 (V) | 26.7 ± 1.9 (D)<br>25.7 ± 1.1 (V) | 897 ± 203 (D)<br>830 ± 175 (V) | Death/BPD 28d |
| Gursoy 2014 | Turkey | 1 | 2006 – 2009<br>(D) 2012 (V) | Cohort | 652 (D)<br>172 (V) | 150 (D)<br>54 (V) | NR | 29.4 ± 1.9 (D)<br>28.9 ± 2.3 (V) | 1218 ± 220 (D)<br>1102 ± 251 (V) | BPD 28d |
| Anand 2015 | USA | 2 | NR | Cohort | 49 (D)<br>46 (V) | 16 (D)<br>NR (V) | NR | NR | NR | BPD 28d |
| Mistry 2020 | Australia | NR | NR | Cohort | NR | NR | NR | NR | NR | BPD (NR) |
| Baud 2021 | France | 21 | 2008 – 2014 | RCT | 523 | 125 | 107 | 26.4 ± 0.8 | 854 ± 170 | Death/BPD 36w |
| **Validation only** |  |  |  |  |  |  |  |  |  |  |
| Fowlie 1998 | UK | 6 | 1988 – 1990 | Cohort | 398 | BPD 28d 75<br>BPD 36w 31 | 81 | 29.8 ± 2.5 | 1065 ± 186 | BPD 28d & BPD 36w |
| Hentschel 1998 | Germany | 1 | 1991 – 1993 | Cohort | 188 | BPD 28d 61<br>BPD 36w 45 | 30 | 28.6 ± 0.3 | 1101 ± 281 | BPD 28d & BPD 36w |
| Schroeder 1998 | Germany | 1 | 1985 – 1992 | Cohort | 103 | 59 | NR | 28.5 ± 1.9 | 1000 ± 153 | BPD 28d |
| Srisuparp 2003 | USA | 1 | 1996 – 1997 | Cohort | 138 | 47 | 24 | 27.6 ± 2.4 | 995 ± 247 | BPD 36w |
| Thowfique 2010 | Singapore | 1 | 2006 – 2007 | Cohort | 388 | 59 | 40 | 28.7 ± 3 | 1029 ± 251 | BPD (NR) |
| Carvalho 2011 | Brazil | 2 | 2002 – 2009 | Cohort | 86 | 20 | 18 | 28.3 ± 1.8 | 851 ± 233 | BPD 36w |
| Onland 2013 | Worldwide | 85 | 1986 – 2004 | 10 RCTs | 3229 | 1094* | 582* | 27.3 ± 3.8 | 989 ± 315 | BPD36w & Death/BPD 36w |
| Truog 2014 | USA | 1 | 2008 – 2010 | Cohort | 158 | 115 | 15 | NR | NR | Death/BPD 36w |
| Sullivan 2016 | USA | 1 | 2004 – 2014 | Cohort | 566 | 98 | 51 | 28.6 ± 2.9 | NR | BPD 36w |
| Ozcan 2017 | Turkey | 1 | 2011 – 2012 | Cohort | 246 | 28 | 32 | 29.2 ± 2.15 | 1323 ± 331 | BPD 36w |
| Gulliver 2018 | USA | 1 | 2010 – 2016 | Cohort | 622 | 223 | 61 | 27.0 ± 1.9 | 963 ± 301 | Death/BPD 36w |
| Vasquez 2018 | Colombia | 2 | 2010 – 2016 | Cohort | 335 | 68 | NR | 31 ± 1.5 | 1328 ± 328 | BPD 28d |
| Jung 2019 | Korea | 1 | 2010 – 2014 | Cohort | 138 | 57 | 17 | 25.9 ± 1.3 | 780 ± 225 | Death/sev BPD 36w |
| Lee 2019 | Korea | 67 | 2013 – 2016 | Cohort | 6938 | 1916 | 957 | 28.3 ± 2.4 | 1059 ± 283 | BPD 36w |
| Baker 2020 | Australia | 2 | 2016 – 2017 | Cohort | 187 | 72 (sev BPD) | 18 | 26.6 ± 1.5 | 872 ± 178 | Death/sev BPD 28d |
| Bhattacharjee 2020 | USA | 1 | 2012 – 2013 | Cohort | 69 | 31 | 5 | 24.75 ± 1.5 | 722 ± 160 | Sev BPD 36w |
| Sotodate 2020 | Japan | 1 | 2010 – 2017 | Cohort | 171 | 74 | 19 | 25.5 ± 1.6 | 741.3 ± 208.2 | BPD 36w |
| Steocklin 2020 | Australia | 1 | 2017 – 2018 | Cohort | 32 | NR | NR | 26.2 ± 1.0 | NR | BPD 36w |
| Alonso 2021 | Spain | 1 | 2013 – 2020 | Cohort | 202 | BPD 28d 58<br>BPD 36w 21 | 23 | 29.5 ± 2.1 | 1142 ± 256 | BPD 28d & BPD 36w |
| Rysavy 2021 | USA | 14 (RCT)<br>32 (Cohort) | 1996 – 1997 (RCT)<br>2016 – 2018 (Cohort) | RCT / Cohort | 807 (RCT)<br>2370 (Cohort) | 356 (RCT)<br>1417 death / BPD (Cohort) | 114 (RCT)<br>1417 death / BPD (Cohort) | 25.8 ± 1.8 (RCT)<br>NR (Cohort) | 770 ± 136 (RCT)<br>NR (Cohort) | Death/BPD 36w |

70% of the 44 derivation studies used logistic regression to develop the BPD prediction tool, with 11% used univariate analysis; 5% used clinical consensus as well as a combination of logistic regression and classification and tree analysis (CART) respectively; and 2% used CART, gradient boosting, Bayesian network and a combination of logistic regression and support vector machine, respectively. Complete case analysis was used in 41% of the included derivation studies, while handling of missing data was not reported in the remaining 59%. Internal and external validation was done in 25% and 30% of the studies, respectively. Validation was not done in the remaining 45% of studies. 75% of the studies assessed discrimination using C-statistics. In contrast, only 16% of the studies evaluated calibration using the goodness of fit (five studies), calibration plot (1 study) and O:E ratio (1 study). Of the 44 models, ten (23%), eight (18%) and four (9%) models provided a formula, score chart and nomogram respectively. Only two (5%) models provided an online calculator (**Table 2**).

**Table 2.** Methodology used by the derivation studies. NR = Not reported. CART = Classification and regression tree. SVM = Support vector machine. ROC = receiver operating characteristics curve. O:E ratio = Observed:expected ratio.

| Study | Model Development |  |  |  |  | Model Validation |  |  |
| --- | --- | --- | --- | --- | --- | --- | --- | --- |
|  | Approach | Continuous | Missing value | Predictor selection | Presentation | Approach | Discrimination | Calibration |
| Cohen 1983 | Clinical consensus | Categorical | Complete case | Clinical consensus | NR | Same cohort | NR | None |
| Palta 1990 | Clinical consensus | Kept linear | NR | Clinical consensus | Formula | Same cohort | NR | None |
| Parker 1992 | Regression | Kept linear | Complete case & Replace mean | Univariate → stepwise | Formula | Bootstrapping / Temporal (year) | NR | O:E ratio |
| Corcoran 1993 | Regression | Categorical | Complete case | Univariate → stepwise | Formula | Random split | NR | None |
| Ryan 1994 | Regression | Kept linear | NR | Univariate → stepwise | Formula | New dataset | ROC | None |
| Gortner 1996 | Regression | Kept linear | NR | Univariate → stepwise | Formula | Same cohort | NR | None |
| Rozycki 1996 | Regression | Categorical | Complete case | Univariate → stepwise | Nomogram | New dataset | NR | None |
| Ryan 1996 | Regression | Kept linear | NR | Univariate → stepwise | Formula | Temporal (year) | ROC | None |
| Romagnoli 1998 | Regression | Categorical | NR | Univariate analysis | Formula | New dataset | ROC | None |
| Yoder 1999 | Regression | Categorical | NR | Univariate analysis | Score chart | New dataset | ROC | None |
| Chien 2002 | Regression | Kept linear | Complete case | NR | NR | Same cohort | ROC | Goodness of fit |
| Groves 2004 | Univariate analysis | Kept linear | NR | Stepwise selection | NR | Same cohort | ROC | None |
| Cunha 2005 | Regression | Categorical | NR | Univariate → stepwise | Score chart | Same cohort | NR | None |
| Kim 2005 | Regression | Categorical | Complete case | Univariate → stepwise | Score chart | New dataset | ROC | None |
| Choi 2006 | Regression | Categorical | NR | Univariate → stepwise | Score chart | Same cohort | ROC | None |
| Henderson-Smart 2006 | Regression | Categorical | Complete case | Univariate → stepwise | Formula | Temporal (year) | ROC | Goodness of fit |
| Bhering 2007 | Regression | Categorical | Complete case | Stepwise selection | Score chart | New dataset | ROC | Goodness of fit |
| May 2007 | Univariate analysis | Kept linear | NR | No selection | NR | New dataset | ROC | None |
| Ambalavanan 2008 | Regression & CART | Kept linear | Complete case | Stepwise selection | Nomogram | Cross-validation | ROC | None |
| Laughon 2011 | Regression | Kept linear | NR | Univariate → stepwise | Formula/Online tool | New dataset | ROC | None |
| Gottipati 2012 | Regression | Kept linear | NR | NR | Score chart | Same cohort | NR | None |
| Roth-Kleiner 2012 | Regression | Categorical | NR | Stepwise selection | Score chart | Cross-validation | ROC | None |
| Chock 2014 | Regression & CART | Kept linear | NR | NR | Nomogram | Same cohort | ROC | None |
| Gursoy 2014 | Regression | Categorical | NR | Univariate → stepwise | Score chart | New dataset | ROC | Goodness of fit |
| Yang 2014 | Univariate analysis | Kept linear | NR | No selection | NR | Same cohort | ROC | None |
| Anand 2015 | Bayesian Network | Categorical | NR | NR | NR | New dataset | ROC | None |
| Ochab 2016 | Regression & SVM | Kept linear | NR | Stepwise selection | NR | Cross-validation | NR | None |
| Wai 2016 | Regression | Kept linear | Complete case | Univariate → stepwise | NR | Same cohort | ROC | None |
| Kim 2017 | Regression | Kept linear | NR | Univariate analysis | NR | Same cohort | NR | None |
| Beltempo 2018 | Regression | Categorical | Complete case | No selection | NR | Same cohort | ROC | None |
| Boghossian 2018 | Regression | Categorical | NR | No selection | NR | Random split | ROC | None |
| Hunt 2018 | Univariate analysis | Categorical | NR | No selection | NR | Same cohort | ROC | None |
| Sullivan 2018 | Regression | Transformation | NR | No selection | NR | Bootstrapping | ROC | None |
| Fairchild 2019 | Regression | Kept linear | NR | Stepwise selection | NR | Same cohort | ROC | None |
| Sun 2019 | Univariate analysis | Kept linear | Complete case | No selection | NR | Same cohort | ROC | None |
| Valenzuela–Stutman 2019 | Regression | Kept linear | NR | Stepwise selection | NR | Random split | ROC | None |
| Dylag 2020 | Regression | Kept linear | Complete case | Stepwise selection | NR | Random split | ROC | None |
| Mistry 2020 | Regression | Kept linear | NR | NR | NR | Same cohort | ROC | None |
| Shah 2020 | Regression | Kept linear | Complete case | A priori knowledge | NR | Same cohort | ROC | None |
| Sharma 2020 | CART | Kept linear | Complete case | CART | Nomogram | Same cohort | ROC | None |
| Vaid 2020 | Gradient boosting | NR | Complete case | NR | NR | Cross-validation | ROC | None |
| Baud 2021 | Regression | Kept linear | NR | Stepwise selection | Formula | Same cohort | ROC | Goodness of fit |
| Shim 2021 | Regression | Kept linear | Complete case | Stepwise selection | Formula | Temporal (year) | NR | None |
| Ushida 2021 | Regression | Kept linear | Complete case | Stepwise selection | Formula/Online tool | Random split | ROC | Calibration |

Of the 53 BPD prediction models identified, 19 used predictors available within 24 hours of age, while 20 and six models relied on predictors available between two to seven days and above seven days of age respectively. Seven models used predictors available at various timepoints while the timepoints were unavailable for one model. The BPD prediction models considered a median of 14 predictors before using a median of five predictors in the final models. The five most used predictors were GA, birthweight, the fraction of inspired oxygen (F_i_O_2_), gender and invasive ventilation requirement, which were used in 33%–69% of models (**Appendix 5**).

#### Risk of Bias

The majority of the studies were assessed to have a low risk of bias for the participants (84%), predictors (92%) and outcome (89%) domains. 60 (94%) studies were assessed to have a high risk of bias in the analysis domain based on the PROBAST tool^13^ due to various reasons including calibration not assessed (55 studies (86%)); small sample size (37 studies (58%)); inappropriate handling of missing data (21 studies (33%)); lack of internal/external validation (9 studies (14%)); inappropriate selection approach for predictors (6 studies (9%)); and inappropriate handling of continuous predictors (2 studies (3%)).

Twenty-one studies (33%) had high applicability concerns in the participant’s domain as they targeted a specific group of very preterm infants, usually infants at a higher risk of BPD (for example, ventilated infants only in 17 studies (27%)). Although universally accessible, predictors used in ten studies (16%) may not be routinely collected. Eight studies (13%) used BPD definitions that deviated against current consensus^10^ (**Figure 2, Appendix 6**).

**Figure 2:**
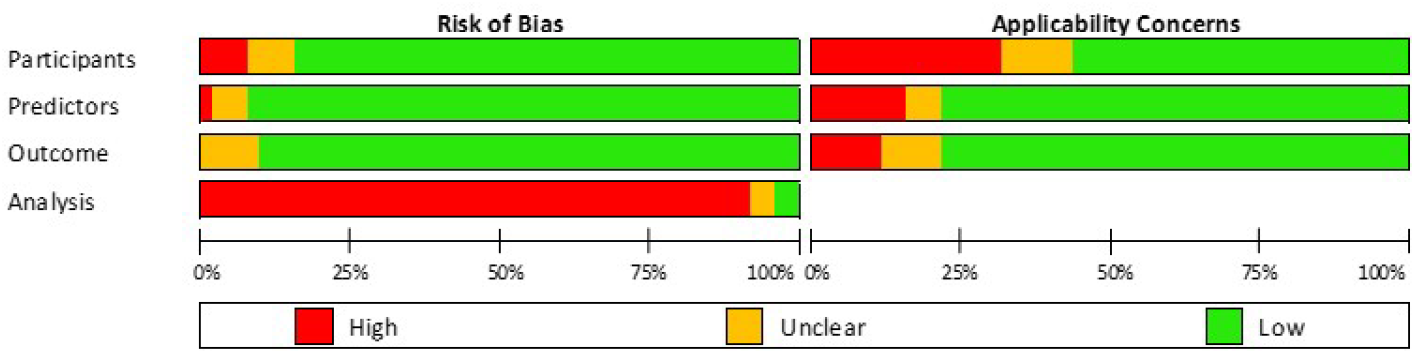
Summary of risk of bias assessments for included studies based on the PROBAST tool^13^.

#### Discrimination

The C-statistics of the included prediction models ranged from 0.52–0.95 in the external validation studies with better performance in models using predictors beyond seven days of age. Meta-analysis could only be done on 22 (50%) and 11 (35%) models for BPD and BPD/death composite outcome respectively, as the remaining 22 and 20 models were only validated in one study. The C-statistics confidence interval (CI) were wide due to the small number of studies in each meta-analysis. The five models with CI above 0.5 for BPD from the meta-analysis were CRIB I^23^, CRIB II^24^ as well as Valenzuela–Stutman 2019^25^ (Birth, Day 3 and 14 models). Similarly, for the BPD/death composite, the five models with CI above 0.5 from the meta-analysis were Laughon 2011^26^ (Day 1, 3, 7 and 14 models) and Valenzuela–Stutman 2019^25^ (Day 14 model) (**Appendix 7**). Meta-analysis for the Valenzuela–Stutman 2019 models^25^ could only be performed after including validation findings from our cohort study.

#### Calibration

The O:E ratio was reported in four external validation studies^27-30^ evaluating six prediction models (Rozycki 1996^27^, Parker 1992^28^ and Laughon 2011^26^ (Day 1, 3, 7 and 14 models)) with considerable variation in the O:E ratio among the included models. Meta-analysis of the O:E ratio could only be done on one model (Laughon 2011^26^ (Day 1)) with an O:E ratio of 0.96 (95% CI 0.85–0.99) (**Appendix 8**).

The calibration plot was reported in three studies^29-31^, assessing six models (Palta 1990^32^, Sinkin 1990^33^, Ryan 1996^34^, Kim 2005^35^ as well as Laughon 2011^26^ (Day 1 and 3 models) (**Appendix 9**).

#### Classification

No studies reported net benefit or decision curve analyses.

#### Heterogeneity and reporting deficiencies

As there were less than ten validation studies in a meta-analysis, subgroup analysis and funnel plots were not performed. Sensitivity analysis was not performed as all studies had an overall high risk of bias except for two studies^29 36^.

#### Summary of findings

Due to the lack of validation studies, a conclusion could only be made for one model Laughon 2011^26^. There was moderate quality of evidence that the discrimination and calibration performances of the Laughon 2011^26^ model in predicting the BPD/death composite outcome using predictors at day one of age with C-statistic of 0.76 (95% CI 0.70–0.81) and O:E ratio of 0.96 (95% CI 0.85–0.99). The evidence was downgraded by one level due to study limitation whereby there was variation in the BPD definition used, as well as some studies recruiting high-risk infants only (such as invasively ventilated infants) to validate the model.

### External validation

#### Patient cohort

After exclusions (**Appendix 10**), 62,864 very preterm infants were included (**Appendix 11**). 17,775 (31%) infants developed BPD while 5,718 (9%) infants died before discharge from the neonatal unit.

#### Model performance

We were able to externally validate six prediction models (Henderson-Smart 2006^37^, Valenzuela–Stutman 2019^25^ (Day1, 3 and 14 models), Shim 2021^38^ and Ushida 2021^39^) in our retrospective cohort. The variables in the remaining models were not available in our cohort. The discrimination (C-statistics) and calibration (O:E ratio and calibration plot) (**Figure 3**) performances were variable among the models. Although the models displayed fair to good discrimination with C-statistics of 0.70–0.90, they had poor calibration as indicated by the calibration plot and O:E ratio between 0.39– 2.31. The Valenzuela–Stutman 2019 models^25^ appear to overestimate the predicted risk, whereas the remaining three models (Henderson-Smart 2006^37^, Shim 2021^38^ and Ushida 2021^39^) tend to underestimate the predicted risk. Of the six externally validated models, four models (Henderson-Smart 2006^37^, Valenzuela–Stutman 2019^25^ (Day 14 models), Shim 2021^38^ and Ushida 2021^39^) indicated superior net benefit across a reasonable range of threshold probabilities of 30% to 60% in deciding postnatal corticosteroid treatment in the decision curve analysis (**Appendix 12**). The threshold probabilities used were identified in a meta-regression of 20 randomised controlled trials^8^.

**Figure 3:**
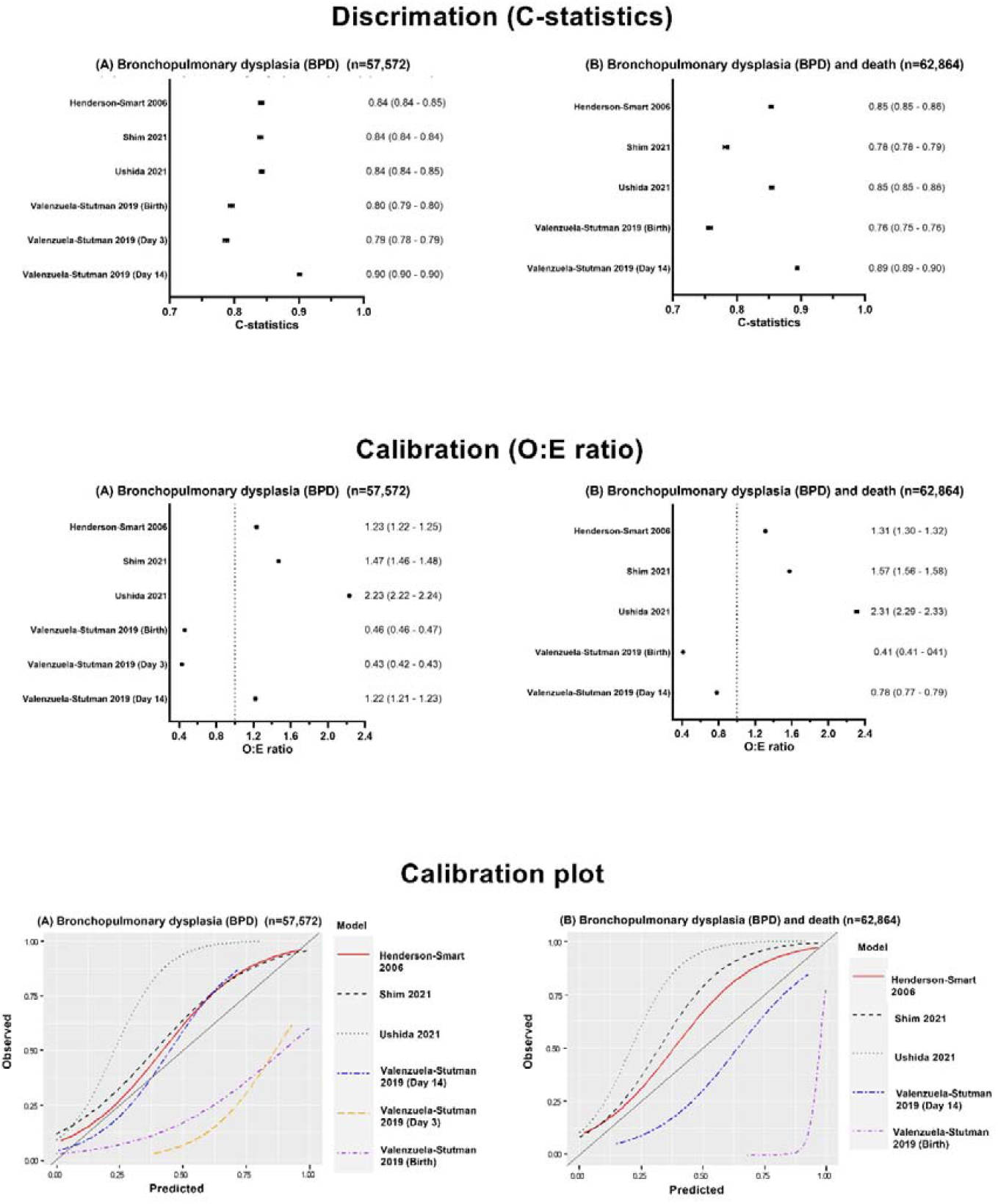
Discrimination (C-statistics) and calibration (O:E ratio and calibration plots) characteristics of prediction models externally validated using a retrospective cohort for (A) bronchopulmonary dysplasia (BPD) (n=57,572) and (B) composite BPD and death (n=62,864).

## Discussion

Our study is an update to the systematic review carried out nearly a decade ago^31^, with a further 27 prediction models identified since the last review. Our systematic review identified 64 studies that developed and/or validated 53 BPD prediction models with meta-analysis carried out on 22 models. Due to the lack of external validation studies, we could not identify a prediction model for routine clinical use. Further external validation, including assessment of both discrimination and calibration performances in a population similar to that whereby the model will be used, is needed before any model could be adopted in clinical practice. However, the most promising prediction model that could be considered based on our meta-analysis was Laughon 2011^26^ in predicting the BPD/death composite outcome using predictors at day one of age. Further re-calibration of the model based on the local population of interest, with re-assessment of its performance in subsequent external validation studies (if re-calibrated), may be needed before being used in clinical practice.

We have also externally validated six prediction models^25 37-39^ in our retrospective population-based cohort study. Although they have fair to good discrimination, they were not well calibrated in our cohort. To be useful, prediction tools need to be generalizable to current datasets highlighting the importance of external validation.

### Implications for clinical practice and research

The implementation of BPD prediction models in clinical practice is limited by the lack of external validation of the published models. Less than a third of the identified prediction models were externally validated. Furthermore, half of the externally validated models were only validated by one study. This potentially limits the generalisability of the model performance to other infant populations.

### Sample size

Most external validation studies had small sample sizes or were restricted to specific high-risk infant populations (for example, ventilated infants only). Furthermore, 61% of studies were single centre only. This potentially limits the generalisability of the models. It is recommended that prediction model development studies should have at least 20 infants with the outcome of interest for each candidate predictor, while validation studies should have at least 100 infants with the outcome^13^.

### Missing values

The majority of the studies did not report missing data or excluded infants with missing data. A clear description of the handling of missing data should be provided. Complete case analysis should be avoided if possible^13^.

### Variation in prediction timepoint and outcome definition

Nearly three-quarters of the included prediction models predicted BPD, the remainder predicting the BPD/death composite outcome. As death and BPD are semi-competitive risks, infants who died before 36 weeks CGA may have a higher risk of developing BPD if they had survived until 36 weeks CGA. Hence, the potential predictive information of death should be accounted for in BPD prediction modelling. The included models also made predictions at a variety of timepoints. Therefore, a meta-analysis of the models was difficult and may limit the clinical settings in which the model can be used. It may be sensible for the performance of future prediction models to be externally validated for BPD as well as the BPD/death composite outcome at three prediction timepoints of one, seven days and 14 days of age. These timepoints would allow timely preventative treatment or research recruitment to be targeted to high-risk infants.

### Predictors

The predictors used in the model should be easily assessed routinely during daily clinical practice and not dependable on clinical practice, such as weight loss and fluid intake. Future prediction models should also be dynamic, accounting for the changing status of the infant over time and clinical trajectory.

Predictor selection based on traditional stepwise approach or univariable analysis should be avoided, especially in small datasets. Instead, predictor selection based on a priori knowledge or statistical approach not based on prior statistical tests between predictor and outcome (e.g. principal component analysis) may be better methods^13^.

### Model performance

Both discrimination (C-statistics) and calibration (calibration plot or O:E ratio) performances of the prediction models need to be assessed during external validation. A model with fair to good discrimination may be poorly calibrated^31^. Hosmer-Lemeshow goodness-of-fit test alone without other calibration measures was found not to be a suitable method to assess calibration as it is sensitive to sample size^13^. The test is often non-significant (i.e. good calibration) in small datasets while usually significant (i.e. poor calibration) in large datasets. Since the recommendation to assess calibration in the last review nearly a decade ago^31^, only two further studies^29 30^ assessed calibration using calibration plots or O:E ratios.

An impact analysis was not carried out in any of the identified prediction models to evaluate if the prediction model improved patient outcomes. Decision curve analysis^40^ may be used as an initial screening method to assess the net benefit of using the prediction model before carrying out further impact analysis. Decision curve analysis can be used on the external validation dataset without further data collection.

### Practicality of model

Prediction models developed should be practical and easy to use at the bedside. Only two published models^26 39^ provided online calculators to allow easy access risk assessment.

Changes in clinical practice and rising BPD rates, potentially makes previously published models outdated affecting its predictive ability. Over half of the published models used data from babies born more than a decade ago. Hence, new models should consider a built-in feature to allow it to learn from future babies and adapt its performance to new practices.

### Strength

The systematic review was carried out based on standard Cochrane methodologies as well as recent recommendations for meta-analysis of prediction models^15^ and risk of bias assessment^13^. There were no language nor date restrictions. The review is anticipated to guide clinicians and researchers in not only developing and/or validating BPD prediction models in very premature infants based on recommendations of the review, but also in identifying the most promising prediction model to be externally validated in their local population.

The use of recent routinely collected clinical information in our external validation study, coupled with its large population coverage, provides an accurate representation of the current neonatal practice in England and Wales. This large cohort of nearly 63,000 very preterm infants, including infants receiving both invasive and non-invasive ventilation, forms an ideal cohort to externally validate and assess BPD prediction models.

### Limitation

Only six out of the 53 identified prediction models could be validated in our cohort. Hence, the performance of the remaining models in our cohort was unclear. However, it is crucial that future models should only use predictors that are easily assessed in clinical practice to ensure their successful clinical implementation.

## Conclusion

As preterm infant survival increases, more survivors are diagnosed with BPD along with the long-term respiratory and neurological consequences. Despite almost a doubling in the number of BPD prediction models published over the last decade, most identified in our systemic review are not used in routine clinical practice. This is due to a lack of good quality external validation studies assessing their performance on the local population of interest. Furthermore, calibration of the models is often not appropriately evaluated in most of the models. Models should be externally validated with a subsequent impact analysis before being adopted in clinical practice. Decision curve analysis may be a good screening tool to assess the net benefit of the tool prior to impact analysis.

Our systematic review has also made recommendations for future BPD prediction models including consideration of additional predictors, a more dynamic model accounting for changes in the infant’s condition over time and their trajectory, and the ability to adapt performance with evolving clinical practice. A good quality, well-validated BPD prediction tool is needed to provide personalised preventative treatment and allow targeted trial recruitment to reduce the long-term impact in this vulnerable and expanding population.

## Data Availability

All data produced in the present study are available upon reasonable request to the authors

## Acknowledgement

We would like to thank Prof Valenzuela–Stutman in providing the algorithm for the Valenzuela–Stutman models^25^. We are grateful to all the families that agreed to include their baby’s data in the NNRD, the health professionals from the UK Neonatal Collaborative (**Appendix 13**) who recorded data and the Neonatal Data Analysis Unit team.

## Funding statement

This research received no specific funding.

## Competing interest statements

None declared.

## Notes

### Competing Interest Statement

The authors have declared no competing interest.

### Clinical Protocols

https://www.crd.york.ac.uk/prospero/display_record.php?ID=CRD42020205215

### Author Declarations

Ethical approval was granted by the Sheffield Research Ethics Committee (REC reference 19/YH/0115).

