## Supplementary Data for "Prediction models of bronchopulmonary dysplasia: a systematic review and meta-analysis with validation"

### Appendix

#### Appendix 1: Search strategy

##### CENTRAL via CRS Web

1. MeSH descriptor: [Risk Assessment] explode all trees

2. MeSH descriptor: [Models, Statistical] explode all trees

3. (((predict* or multicomponent or multivariable) near model*) or (predict* near (outcome* or risk* or model*)) or ((history or variable* or criteria or scor* or characteristic* or finding* or factor* or value*) near (predict* or model* or decision* or identif* or prognos*)) or (decision* near (model* or clinical* or logistic model*)) or (prognostic near (history or variable* or criteria or scor* or characteristic* or finding* or factor* or model*)) or (observ* near (variation or model*)))

4. (#1 or #2 or #3) not pain

5. MeSH descriptor: [Bronchopulmonary Dysplasia] explode all trees

6. bronchopulmonary dysplasia or BPD or chronic lung disease or CLD

7. #5 or #6

8. MESH DESCRIPTOR Infant, Newborn EXPLODE ALL AND CENTRAL:TARGET

9. infant or infants or infant's or "infant s" or infantile or infancy or newborn* or "new born" or "new borns" or "newly born" or neonat* or baby* or babies or premature or prematures or prematurity or preterm or preterms or "pre term" or premies or "low birth weight" or "low birthweight" or VLBW or LBW or ELBW or NICU AND CENTRAL:TARGET

10. #8 or #9

11. #4 and #7 and #10

##### Medline via Ovid - Ovid MEDLINE(R) and Epub Ahead of Print, In-Process & Other Non-Indexed Citations, Daily and Versions(R):

1. (Risk Assessment/ or Models, Statistical/ or (predict* or validat* or rule* or scor*).ti,ab. or ((predict* or multicomponent or multivariable) adj3 model*).mp. or (predict* adj5 (outcome* or risk* or model*)).ti,ab. or ((history or variable* or criteria or scor* or characteristic* or finding* or factor* or value*) adj5 (predict* or model* or decision* or identif* or prognos*)).ti,ab. or (decision* adj5 (model* or clinical* or logistic model*)).ti,ab. or (prognostic adj5 (history or variable* or criteria or scor* or characteristic* or finding* or factor* or model*)).ti,ab. or (observ* adj3 (variation or model*)).ti,ab.) not pain.mp.

2. bronchopulmonary dysplasia or BPD or chronic lung disease or CLD

3. #1 and #2

4. exp infant, newborn/

5. (newborn* or new born or new borns or newly born or baby* or babies or premature or prematurity or preterm or pre term or low birth weight or low birthweight or VLBW or LBW or infant or infants or 'infant s' or infant's or infantile or infancy or neonat*).ti,ab.

6. #4 or #5

7. #3 and #6

##### Cumulative Index to Nursing and Allied Health Literature (CINAHL) via EBSCOhost

1. ((MM “Risk Assessment”) OR (MM “Models, Statistical”) or TI ((predict* or validat* or rule* or scor*) or ((predict* or multicomponent or multivariable) N3 model*) or (predict* N3 (outcome* or risk* or model*)) or ((history or variable* or criteria or scor* or characteristic* or finding* or factor* or value*) N3 (predict* or model* or decision* or identif* or prognos*)) or (decision* N3 (model* or clinical* or logistic model*)) or (prognostic N3 (history or variable* or criteria or scor* or characteristic* or finding* or factor* or model*)) or (observ* N3 (variation or model*))) or AB ((predict* or validat* or rule* or scor*) or ((predict* or multicomponent or multivariable) N3 model*) or (predict* N3 (outcome* or risk* or model*)) or ((history or variable* or criteria or scor* or characteristic* or finding* or factor* or value*) N3 (predict* or model* or decision* or identif* or prognos*)) or (decision* N3 (model* or clinical* or logistic model*)) or (prognostic N3 (history or variable* or criteria or scor* or characteristic* or finding* or factor* or model*)) or (observ* N3 (variation or model*)))) NOT TX pain

2. MM "Bronchopulmonary Dysplasia" OR (bronchopulmonary dysplasia or BPD or chronic lung disease or CLD)

3. #1 AND #2

4. infant or infants or infant's or infantile or infancy or newborn* or "new born" or "new borns" or "newly born" or neonat* or baby* or babies or premature or prematures or prematurity or preterm or preterms or "pre term" or premies or "low birth weight" or "low birthweight" or VLBW or LBW

5. #3 AND #4

*Searching other resources*

Reference lists of included studies and previous reviews were examined alongside proceedings of annual meetings of the Paediatric American Societies (from 1993 onwards), the European Society for Paediatric Research (from 1995 onwards), the Royal College of Paediatrics and Child Health (from 2000 onwards), and the Perinatal Society of Australia and New Zealand (from 2000 onwards). Studies reported as abstracts were included if sufficient information was obtained.

#### Appendix 2: Preliminary study selection, data extraction and risk of bias forms

##### 2a Data extraction spreadsheet (adapted from the CHARMS checklist^41^ and Cochrane exemplar by Pace et al^42^)

| Author ID: |  | Date of extraction: |  |
| --- | --- | --- | --- |
| **Domain** | **Key items** | **Extracted Data** | **Reported on page number** |
| Study citation | Authors, journal name, year, volume, pages |  |  |
| Source of data | e.g. cohort (prospective or retrospective), case-control, randomised trial participants |  |  |
| Participants | Participant eligibility and recruitment method (e.g. consecutive participants, location(s) / country(s), number of centres, setting) |  |  |
|  | Inclusion criteria |  |  |
|  | Exclusion criteria |  |  |
|  | Study dates/period of data collection |  |  |
|  | Participant characteristics |  |  |
|  | Gestational age at birth |  |  |
|  | Birthweight |  |  |
|  | Gender |  |  |
|  | Ethnicity |  |  |
|  | Antenatal corticosteroids use |  |  |
|  | Surfactant |  |  |
|  | Number of non-ventilated infants |  |  |
|  | Duration of invasive ventilation |  |  |
|  | Postnatal corticosteroids |  |  |
| Outcome(s) to be predicted | Definition of BPD used (e.g. respiratory support requirement at 28 days old or 36 weeks CGA) |  |  |
|  | Was the physiological challenge of oxygen withdrawal performed? |  |  |
|  | Was the same BPD definition and method of assessment used for all participants? |  |  |
|  | Type of outcome (e.g. BPD or composite BPD and death before discharge from neonatal units) |  |  |
|  | Was the outcome assessed without knowledge of candidate predictors (i.e. blinded)? |  |  |
|  | Was candidate predictors part of the outcome? |  |  |
| Candidate predictors | Number of predictors |  |  |
|  | Definition and method of measurement for each candidate predictors |  |  |
|  | Age of infants/timing of predictor measurement |  |  |
|  | Were predictors assessed blinded for the outcome, and for each other (if relevant)? |  |  |
|  | Handling of continuous predictors in the modelling (e.g. linear, non-linear transformation categorised) |  |  |
| Sample size | Number of participants |  |  |
|  | Number of events |  |  |
|  | Number of events per number of candidate predictor |  |  |
| Missing data | Number of participants with any missing value (including predictors and outcomes) |  |  |
|  | Number of participants with missing data for outcome |  |  |
|  | Number of participants with missing data for each predictor |  |  |
|  | Handling of missing data (e.g. complete case analysis, imputation) |  |  |
| Model development | Modelling method (e.g. logistic regression, neural network) and statistical software used |  |  |
|  | Modelling assumptions satisfied |  |  |
|  | Method for selection of predictors **for inclusion in** multivariable modelling (e.g. all candidate predictors, pre-selection based on unadjusted association with the outcome) |  |  |
|  | Method for selection of predictors **during** multivariable modelling (e.g. full model approach, backward or forward selection) and criteria used (e.g. p-value, Akaike Information Criteria) |  |  |
|  | Shrinkage of predictor weights or regression coefficients (e.g. no shrinkage, uniform shrinkage, penalised estimation) |  |  |
| Model performance | Discrimination measure (C-statistics) with confidence interval or standard error |  |  |
|  | Calibration measure (O:E ratio, calibration plot or slope, Hosmer-Lemeshow test) with confidence interval or standard error |  |  |
|  | Classification measure (e.g. net benefit analysis, sensitivity, specificity, positive predictive value, negative predictive value) and whether a priori cut-offs were used |  |  |
| Model evaluation | Method used for testing model performance: (a) Developmental dataset only (random split, re-sampling methods (e.g. bootstrap), (b) Separate external validation (e.g. temporal, geographical) |  |  |
|  | If poor validation, was model adjusted or updated (e.g. intercept re-calibrated, predictor effects adjusted, new predictors added)? |  |  |
| Results | Final model presented including predictor weights |  |  |
|  | Alternative presentation of the model (e.g. nomogram, score chart) |  |  |
|  | Comparison of the distribution of predictors (including missing data) for development and validation datasets |  |  |
| Interpretation and discussion | Interpretation of model (e.g. confirmatory - model useful for practice; exploratory - more research needed) |  |  |
|  | Comparison with other studies (e.g. generalisability, strengths and limitations) |  |  |
|  | Ease of use of the model |  |  |
| Author correspondence | Contact details of the corresponding author |  |  |
| References | Reference of relevant studies cited in the reference list |  |  |

##### 2b PROBAST tool (adapted from Wolff et al^13^ and Moons et al^43^)

**Step 1: Specify your systematic review question**

| **Criteria** | **Specify your systematic review question** |
| --- | --- |
| Intended use of the model: | Prognostic model to predict the occurrence of bronchopulmonary dysplasia at 28 days of age or 36 weeks of corrected gestational age. |
| **Participants** including selection criteria and setting: | Very preterm infants born below 32 weeks of gestational age |
| **Predictors** (used in prediction modelling), including types of predictors (e.g. history, clinical examination, biochemical markers, imaging tests), time of measurement, specific measurement issues (e.g., any requirements/ prohibitions for specialised equipment): | Universally accessible predictors that are available within the first two weeks of age are included.  Non-universally accessible predictors, such as pulmonary function testing, ultrasonography and biomarkers, are excluded. |
| Outcome to be predicted: | Bronchopulmonary dysplasia at 28 days of age or 36 weeks of corrected gestational age |

**Step 2: Classify the type of prediction model evaluation**

| **Classify the evaluation based on its aim** | | | |
| --- | --- | --- | --- |
| **Type of prediction study** | **PROBAST boxes to complete** | **Tick as appropriate** | **Definition for the type of prediction model study** |
| Development only | Development |  | Prediction model development without external validation. These studies may include internal validation methods, such as bootstrapping and cross-validation techniques. |
| Development and validation | Development and validation |  | Prediction model development combined with external validation in other participants in the same article. |
| Validation only | Validation |  | External validation of existing (previously developed) model in other participants. |

This table should be completed once for each publication being assessed and for each relevant outcome in your review.

| Publication reference |
| --- |
| Models of interest |
| Outcome of interest |

**Step 3: Assess the risk of bias and applicability**

PROBAST is structured into four key domains. Each domain is judged for risk of bias (low, high or unclear) and includes signalling questions to help make judgements. Signalling questions are rated as yes (Y), probably yes (PY), probably no (PN), no (N) or no information (NI). All signalling questions are phrased so that "yes" indicates the absence of bias. Any signalling question rated as "no" or "probably no" flags the potential for bias; you will need to use your judgement to determine whether the domain should be rated as "high", "low" or "unclear" risk of bias. The guidance document contains further instructions and examples on rating signalling questions and risk of bias for each domain.

The first three domains are also rated for concerns regarding applicability (low/ high/ unclear) to your review question defined above.

Complete all domains separately for each evaluation of a distinct model. Shaded boxes indicate where signalling questions do not apply and should not be answered.

| **DOMAIN 1: Participants** | | | |
| --- | --- | --- | --- |
| **A. Risk of Bias** | | | |
| Describe the sources of data and criteria for participant selection: | | | |
|  | | Dev | Val |
| 1.1 Were appropriate data sources used, e.g. cohort, RCT or nested case-control study data? | |  |  |
| 1.2 Were all inclusions and exclusions of participants appropriate? | |  |  |
| Risk of bias introduced by the selection of participants | RISK:  (low/ high/ unclear) |  |  |
| Rationale of bias rating: | | | |
| **B. Applicability** | | | |
| Describe included participants, setting and dates: | | | |
| Concern that the included participants and setting do not match the review question | CONCERN:  (low/ high/ unclear) |  |  |
| Rationale of applicability rating: | | | |

| **DOMAIN 2: Predictors** | | | |
| --- | --- | --- | --- |
| **A. Risk of Bias** | | | |
| List and describe predictors included in the final model, e.g. definition and timing of assessment: | | | |
|  | | Dev | Val |
| 2.1 Were predictors defined and assessed in a similar way for all participants? | |  |  |
| 2.2 Were predictor assessments made without knowledge of outcome data? | |  |  |
| 2.3 Are all predictors available at the time the model is intended to be used? | |  |  |
| Risk of bias introduced by predictors or their assessment | RISK:  (low/ high/ unclear) |  |  |
| Rationale of bias rating: | | | |
| **B. Applicability** | | | |
| Concern that the definition, assessment or timing of predictors in the model do not match the review question | CONCERN:  (low/ high/ unclear) |  |  |
| Rationale of applicability rating: | | | |

| **DOMAIN 3: Outcome** | | | | | |
| --- | --- | --- | --- | --- | --- |
| **A. Risk of Bias** | | | | | |
| Describe the outcome, how it was defined and determined, and the time interval between predictor assessment and outcome determination: | | | | | |
|  | | | Dev | | Val |
| 3.1 Was the outcome determined appropriately? | | |  | |  |
| 3.2 Was a pre-specified or standard outcome definition used? | | |  | |  |
| 3.3 Were predictors excluded from the outcome definition? | | |  | |  |
| 3.4 Was the outcome defined and determined in a similar way for all participants? | | |  | |  |
| 3.5 Was the outcome determined without knowledge of predictor information? | | |  | |  |
| 3.6 Was the time interval between predictor assessment and outcome determination appropriate? | | |  | |  |
| Risk of bias introduced by the outcome or its determination | RISK:  (low/ high/ unclear) | |  | |  |
| Rationale of bias rating: | | | | | |
| **B. Applicability** | | | | | |
| At what time point was the outcome determined:  If a composite outcome was used, describe the relative frequency/distribution of each contributing outcome: | | | | | |
| Concern that the outcome, its definition, timing or determination do not match the review question | | CONCERN:  (low/ high/ unclear) | |  | |
| Rationale of applicability rating: | | | | | |

| **DOMAIN 4: Analysis** | | | |
| --- | --- | --- | --- |
| **Risk of Bias** | | | |
| Describe numbers of participants, number of candidate predictors, outcome events and events per candidate predictor: | | | |
| Describe how the model was developed (for example in regards to modelling technique (e.g. survival or logistic modelling), predictor selection, and risk group definition): | | | |
| Describe whether and how the model was validated, either internally (e.g. bootstrapping, cross-validation, random split sample) or externally (e.g. temporal validation, geographical validation, different setting, different type of participants): | | | |
| Describe the performance measures of the model, e.g. (re)calibration, discrimination, (re)classification, net benefit, and whether they were adjusted for optimism: | | | |
| Describe any participants who were excluded from the analysis: | | | |
| Describe missing data on predictors and outcomes as well as methods used for missing data: | | | |
|  | | Dev | Val |
| 4.1 Were there a reasonable number of participants with the outcome? | |  |  |
| 4.2 Were continuous and categorical predictors handled appropriately? | |  |  |
| 4.3 Were all enrolled participants included in the analysis? | |  |  |
| 4.4 Were participants with missing data handled appropriately? | |  |  |
| 4.5 Was the selection of predictors based on univariable analysis avoided? | |  |  |
| 4.6 Were complexities in the data (e.g. censoring, competing risks, sampling of controls) accounted for appropriately? | |  |  |
| 4.7 Were relevant model performance measures evaluated appropriately? | |  |  |
| 4.8 Were model overfitting and optimism in model performance accounted for? | |  |  |
| 4.9 Do predictors and their assigned weights in the final model correspond to the results from the multivariable analysis? | |  |  |
| Risk of bias introduced by the analysis | RISK:  (low/ high/ unclear) |  |  |
| Rationale of bias rating: | | | |

**Step 4: Overall assessment**

Use the following tables to reach overall judgements about risk of bias and concerns regarding the applicability of the prediction model evaluation (development and/or validation) across all assessed domains. *Complete for each evaluation of a distinct model.*

| **Reaching an overall judgement about the risk of bias of the prediction model evaluation** | |
| --- | --- |
| Low risk of bias | If all domains were rated low risk of bias.  If a prediction model was developed without any external validation, and it was rated as low risk of bias for all domains, consider downgrading to **high risk of bias**. Such a model can only be considered as low risk of bias, if the development was based on a very large data set and included some form of internal validation. |
| High risk of bias | If at least one domain is judged to be at **high risk of bias**. |
| Unclear risk of bias | If an unclear risk of bias was noted in at least one domain and it was low risk for all other domains. |

| **Reaching an overall judgement about the applicability of the prediction model evaluation** | |
| --- | --- |
| Low concerns regarding applicability | If low concerns regarding applicability for all domains, the prediction model evaluation is judged to have **low concerns regarding applicability**. |
| High concerns regarding applicability | If high concerns regarding applicability for at least one domain, the prediction model evaluation is judged to have **high concerns regarding applicability**. |
| Unclear concerns regarding applicability | If unclear concerns (but no "high concern") regarding applicability for at least one domain, the prediction model evaluation is judged to have **unclear concerns regarding applicability** overall. |

| **Overall judgement about risk of bias and applicability of the prediction model evaluation** | |
| --- | --- |
| Overall judgement of risk of bias | RISK:  (low/ high/ unclear) |
| Summary of sources of potential bias: | |
| Overall judgement of applicability | CONCERN:  (low/ high/ unclear) |
| Summary of applicability concerns: | |

#### Appendix 3: Definition of predictor and outcome in the external validation cohort study

BPD was defined as respiratory support or oxygen requirement over a three-day period at 36 weeks of corrected gestational age (CGA). Respiratory support at discharge was used if infants were discharged from neonatal units before 36 weeks CGA. Information on room air challenge test was not available. A composite secondary outcome of BPD and death before 36 weeks CGA was also used.

Birthweight centile was calculated using the UK WHO growth chart, with small for gestational age defined as below the 10^th^ birthweight centile. Antenatal steroids were defined as any antenatal steroids received by the mother. Respiratory distress syndrome, maternal hypertension, prolonged rupture of membrane and chorioamnionitis were defined based on the diagnosis entered on admission or discharge.

#### Appendix 4: Reference list of included full-text articles

Cohen A, Taeusch Jr HW. Prediction of risk of bronchopulmonary dysplasia. American Journal of Perinatology 1983: 1(1): 21-22.

Palta M, Gabbert D, Fryback D, Widjaja I, Peters ME, Farrell P, Johnson J. Development and validation of an index for scoring baseline respiratory disease in the very low birth weight neonate. Pediatrics 1990: 86(5): 714-721.

Parker RA, Lindstrom DP, Cotton RB. Improved survival accounts for most, but not all, of the increase in bronchopulmonary dysplasia. Pediatrics 1992: 90(5): 663-668.

Corcoran JD, Patterson CC, Thomas PS, Halliday HL. Reduction in the risk of bronchopulmonary dysplasia from 1980-1990: results of a multivariate logistic regression analysis. European Journal of Pediatrics 1993: 152(8): 677-681.

Ryan SW, Wild NJ, Arthur RJ, Shaw BN. Prediction of chronic neonatal lung disease in very low birthweight neonates using clinical and radiological variables. Arch Dis Child Fetal Neonatal Ed 1994: 71(1): F36-39.

Gortner L, Schroeder P, Schaible T, Moller J, Teichert HM. Bronchopulmonary dysplasia in preterm infants. An eight year retrospective and analysis of risk factors. Monatsschrift Kinderheilkunde 1996: 144(9): 895-900.

Rozycki HJ, Narla L. Early versus late identification of infants at high risk of developing moderate to severe bronchopulmonary dysplasia. Pediatric Pulmonology 1996: 21(6): 345-352.

Ryan SW, Nycyk J, Shaw BN. Prediction of chronic neonatal lung disease on day 4 of life. European Journal of Pediatrics 1996: 155(8): 668-671.

Fowlie PW, Gould CR, Tarnow-Mordi WO, Strang D. Measurement properties of the Clinical Risk Index for Babies - Reliability, validity beyond the first 12 hours, and responsiveness over 7 days. Critical Care Medicine 1998: 26(1): 163-168.

Hentschel J, Friedel C, Maier RE, Bassir C, Obladen M. Predicting chronic lung disease in very low birthweight infants: Comparison of 3 scores. Journal of Perinatal Medicine 1998: 26(5): 378-383.

Romagnoli C, Zecca E, Tortorolo L, Vento G, Tortorolo G. A scoring system to predict the evolution of respiratory distress syndrome into chronic lung disease in preterm infants. Intensive Care Medicine 1998: 24(5): 476-480.

Schroeder P, Gortner L. Predicting bronchopulmonary dysplasia. Intensive Care Medicine 1999: 25(2): 241-242.

Yoder BA, Anwar MU, Clark RH. Early prediction of neonatal chronic lung disease: a comparison of three scoring methods. Pediatric Pulmonology 1999: 27(6): 388-394.

Chien LY, Whyte R, Thiessen P, Walker R, Brabyn D, Lee SK. SNAP-II predicts severe intraventricular hemorrhage and chronic lung disease in the neonatal intensive care unit. Journal of Perinatology 2002: 22(1): 26-30.

Srisuparp P, Marks JD, Khoshnood B, Schreiber MD. Predictive power of initial severity of pulmonary disease for subsequent development of bronchopulmonary dysplasia. Biology of the Neonate 2003: 84(1): 31-36.

Groves AM, Briggs KA, Kuschel CA, Harding JE. Predictors of chronic lung disease in the ‘CPAP era’. Journal of paediatrics and child health 2004: 40(5‐6): 290-294.

Cunha GS, Mezzacappa-Filho F, Ribeiro JD. Risk factors for bronchopulmonary dysplasia in very low birth weight newborns treated with mechanical ventilation in the first week of life. J Trop Pediatr 2005: 51(6): 334-340.

Kim YD, Kim EAR, Kim KS, Pi SY, Kang WC. Scoring method for early prediction of neonatal chronic lung disease using modified respiratory parameters. Journal of Korean Medical Science 2005: 20(3): 397-401.

Choi EN, Ramgung R, Koo HK. Early prediction of Bronchopulmonary Dysplasia (BPD) in Very Low Birth Weight Infants with Mechanical Ventilation in the First Week of Life In: Paediatric Academic Society; 2006; 2006.

Henderson-Smart DJ, Hutchinson JL, Donoghue DA, Evans NJ, Simpson JM, Wright I. Prenatal predictors of chronic lung disease in very preterm infants. Archives of Disease in Childhood -- Fetal & Neonatal Edition 2006: 91(1): F40-45.

Bhering CA, Mochdece CC, Moreira MEL, Rocco JR, Sant'Anna GM. Bronchopulmonary dysplasia prediction model for 7-day-old infants. Jornal De Pediatria 2007: 83(2): 163-170.

May C, Kavvadia V, Dimitriou G, Greenough A. A scoring system to predict chronic oxygen dependency. Eur J Pediatr 2007: 166(3): 235-240.

Ambalavanan N, Van Meurs KP, Perritt R, Carlo WA, Ehrenkranz RA, Stevenson DK, Lemons JA, Poole WK, Higgins RD. Predictors of death or bronchopulmonary dysplasia in preterm infants with respiratory failure. Journal of Perinatology 2008: 28(6): 420-426.

Thowfique I, Agarwal P, Bhavani S, Rajadurai VS. Clinical risk index of babies (CRIB) score and neonatal outcome in very low birthweight (VLBW) infants. Proceedings of Singapore Healthcare 2010: 19(SUPPL. 2): S104.

Carvalho PRN, Moreira MEL, Sá RAM, Lopes LM. SNAPPE-II application in newborns with very low birth weight: Evaluation of adverse outcomes in severe placental dysfunction. Journal of Perinatal Medicine 2011: 39(3): 343-347.

Laughon MM, Langer JC, Bose CL, Smith PB, Ambalavanan N, Kennedy KA, Stoll BJ, Buchter S, Laptook AR, Ehrenkranz RA, Cotten CM, Wilson-Costello DE, Shankaran S, Van Meurs KP, Davis AS, Gantz MG, Finer NN, Yoder BA, Faix RG, Carlo WA. Prediction of bronchopulmonary dysplasia by postnatal age in extremely premature infants. American Journal of Respiratory & Critical Care Medicine 2011: 183(12): 1715-1722.

Gottipati V, McCoy M, Anderson M, Sekar K. A predictive model for describing bronchopulmonary dysplasia outcome in infants requiring intubation and surfactant. Journal of Investigative Medicine 2012: 60(1): 421-422.

Roth-Kleiner M, Chnayna J, Giannoni E, Faouzi M. 596 Early Prediction of Bronchopulmonary Dysplasia (Bpd) by an Easily Available Risk Score. Archives of Disease in Childhood 2012: 97(Suppl 2): A173-A173.

Gursoy T, Derin H, Hayran M, Ovali F. A clinical scoring system to predict the development of bronchopulmonary dysplasia in very low birth weight infants. Journal of Perinatal Medicine 2013: 41(SUPPL. 1).

Ozcan B, Kavurt S, Aydemir O, Gencturk Z, Bas AY, Demirel N. Snappe-II and risk of neonatal morbidities in very low birth weight preterm infants. Intensive Care Medicine 2013: 39(SUPPL. 1): S69-S70.

Onland W, Debray TP, Laughon MM, Miedema M, Cools F, Askie LM, Asselin JM, Calvert SA, Courtney SE, Dani C, Durand DJ, Marlow N, Peacock JL, Pillow JJ, Soll RF, Thome UH, Truffert P, Schreiber MD, Van Reempts P, Vendettuoli V, Vento G, van Kaam AH, Moons KG, Offringa M. Clinical prediction models for bronchopulmonary dysplasia: a systematic review and external validation study. BMC Pediatr 2013: 13: 207.

Li YH, Yan J, Li MX, Xiao ZH, Zhu XP, Pan J, Li XZ, Feng X. Addition of SNAP to perinatal risk factors improves the prediction of bronchopulmonary dysplasia or death in critically ill preterm infants. Bmc Pediatrics 2013: 13: 9.

Gursoy T, Hayran M, Derin H, Ovali F. A clinical scoring system to predict the development of bronchopulmonary dysplasia. American Journal of Perinatology 2014: 32(7): 659-665.

Gursoy T, Hayran M, Derin H, Ovali F. Prediction of the development of bronchopulmonary dysplasia preterm infants: A new scoring system and prospective validation. Journal of Maternal-Fetal and Neonatal Medicine 2014: 27(SUPPL. 1): 210-211.

Chock VY, Punn R, Oza A, Benitz WE, Van Meurs KP, Whittemore AS, Behzadian F, Silverman NH. Predictors of bronchopulmonary dysplasia or death in premature infants with a patent ductus arteriosus. Pediatr Res 2014: 75(4): 570-575.

Truog WE, Nyp MF, Taylor J, Gratny LL, Escobar H, Manimtim WM, Lachica CI, Khmour A, Oluola OO, Oshodi AA, Norberg M, Dai H, Pallotto EK. Infants born at <29 weeks: pulmonary outcomes from a hybrid perinatal system. Journal of perinatology : official journal of the California Perinatal Association 2014: 34(1): 59-63.

Yang JY, Cha J, Shim SY, Cho SJ, Park EA. The relationship between eosinophilia and bronchopulmonary dysplasia in premature infants at less than 34 weeks' gestation. Korean Journal of Pediatrics 2014: 57(4): 171-177.

Anand V, Schneeberger D, Piedimonte G. A probabilistic model for prediction of bronchopulmonary dysplasia in pre-term and term babies admitted to the NICU. American Journal of Respiratory and Critical Care Medicine 2015: 191(MeetingAbstracts).

Jang JH, Jung YH, Kimn HS, Kim EK, Shin SH. The effectiveness of respiratory severity score for prediction of severe bronchopulmonary dysplasia or mortality in preterm infants. European Journal of Pediatrics 2016: 175(11): 1463.

Ochab M, Wajs W. Expert system supporting an early prediction of the bronchopulmonary dysplasia. Computers in Biology & Medicine 2016: 69: 236-244.

Sullivan BA, McClure C, Hicks J, Lake DE, Moorman JR, Fairchild KD. Early Heart Rate Characteristics Predict Death and Morbidities in Preterm Infants. Journal of Pediatrics 2016: 174: 57-62.

Wai K, Kohn MA, Ballard R, Black D, Keller RL. Early cumulative supplemental oxygen (O2) exposure predicts bronchopulmonary dysplasia (BPD) in extremely low gestational age newborns (ELGAN). Journal of Investigative Medicine 2016: 64(1): 336.

Kim HR, Kim JY, Yun BL, Lee B, Choi CW, Kim BI. Interstitial pneumonia pattern on day 7 chest radiograph predicts bronchopulmonary dysplasia in preterm infants. BMC Pediatrics 2017: 17(1): 125.

Ozcan B, Kavurt AS, Aydemir O, Gencturk Z, Bas AY, Demirel N. SNAPPE-II and risk of neonatal morbidities in very low birth weight preterm infants. Turkish Journal of Pediatrics 2017: 59(2): 105-112.

Boghossian NS, Geraci M, Edwards EM, Horbar JD. Neonatal and fetal growth charts to identify preterm infants <30 weeks gestation at risk of adverse outcomes. American Journal of Obstetrics and Gynecology 2018: 219(2): 195.

Gulliver K, Yoder BA. Bronchopulmonary dysplasia: effect of altitude correction and role for the Neonatal Research Network Prediction Algorithm. Journal of Perinatology 2018: 38(8): 1046-1050.

Hunt KA, Dassios T, Ali K, Greenough A. Prediction of bronchopulmonary dysplasia development. Archives of Disease in Childhood-Fetal and Neonatal Edition 2018: 103(6): F598-+.

Sullivan BA, Wallman-Stokes A, Isler J, Sahni R, Moorman JR, Fairchild KD, Lake DE. Early Pulse Oximetry Data Improves Prediction of Death and Adverse Outcomes in a Two-Center Cohort of Very Low Birth Weight Infants. American Journal of Perinatology 2018: 35(13): 1331-1338.

Vasquez P, Romero Diaz HA, Gomez MJ, Leal L, Yejas Navarro LM. Bronchopulmonary dysplasia predictor scale validation in preterm newborns in two neonatal units at 2600m above sea level. Infant Behavior & Development 2018: 52: 1-8.

Baker EK, Davis PG. BPD outcome estimator: Applicability in preterm infants 2018. Journal of Paediatrics and Child Health 2019: 55(Supplement 1): 7.

Gulliver K, Yoder B. Does the NRN BPD prediction algorithm correctly identify BPD risk using different BPD definitions at altitude? Journal of Investigative Medicine 2019: 67(1): 121.

Beltempo M, Shah PS, Ye XY, Afifi J, Lee S, McMillan DD. SNAP-II for prediction of mortality and morbidity in extremely preterm infants. J Matern Fetal Neonatal Med 2019: 32(16): 2694-2701.

Fairchild KD, Nagraj VP, Sullivan BA, Moorman JR, Lake DE. Oxygen desaturations in the early neonatal period predict development of bronchopulmonary dysplasia. Pediatr Res 2019: 85(7): 987-993.

Jung YH, Jang J, Kim HS, Shin SH, Choi CW, Kim EK, Kim BI. Respiratory severity score as a predictive factor for severe bronchopulmonary dysplasia or death in extremely preterm infants. BMC Pediatr 2019: 19(1): 121.

Lee SM, Lee MH, Chang YS. The Clinical Risk Index for Babies II for Prediction of Time-Dependent Mortality and Short-Term Morbidities in Very Low Birth Weight Infants. Neonatology 2019: 116(3): 244-251.

Sun YY, Chen CE, Zhang XX, Weng XC, Sheng AQ, Zhu YK, Chen SJ, Zheng XX, Lu CS. High Neutrophil-to-Lymphocyte Ratio Is an Early Predictor of Bronchopulmonary Dysplasia. Frontiers in Pediatrics 2019: 7: 9.

Valenzuela-Stutman D, Marshall G, Tapia JL, Mariani G, Bancalari A, Gonzalez Á. Bronchopulmonary dysplasia: risk prediction models for very-low- birth-weight infants. J Perinatol 2019: 39(9): 1275-1281.

Baker EK, Davis PG. Bronchopulmonary Dysplasia Outcome Estimator in Current Neonatal Practice. Acta Paediatr 2020.

Bhattacharjee I, Das A, Collin M, Aly H. Predicting outcomes of mechanically ventilated premature infants using respiratory severity score. Journal of Maternal-Fetal and Neonatal Medicine 2020.

Mistry AE, De Waal K. Respiratory severity score: Predicting bronchopulmonary dysplasia in preterm infants. Journal of Paediatrics and Child Health 2020: 56(SUPPL 1): 105.

Pillow Jane J, Benjamin S, Harshad P, Jane C, Pillow Jane J. Bronchopulmonary dysplasia can be predicted by early assessment of shift of the SPO2 vs. PIO2 curve in extremely preterm infants. Journal of Paediatrics and Child Health 2020: 56(SUPPL 1): 46-47.

Shah SI, Aboudi D, La Gamma EF, Brumberg HL. Respiratory Severity Score greater than or equal to 2 at birth is associated with an increased risk of mortality in infants with birth weights less than or equal to 1250 g. Pediatric Pulmonology 2020: 55(12): 3304-3311.

Vaid A, Padilla L, Rehan V. Prediction of bronchopulmonary dysplasia on postnatal day one by gradient boosting, an ensemble machine learning algorithm. Journal of Investigative Medicine 2020: 68(1): A121.

Dylag AM, Kopin HG, O'Reilly MA, Wang HY, Davis SD, Ren CL, Pryhuber GS. Early Neonatal Oxygen Exposure Predicts Pulmonary Morbidity and Functional Deficits at 1 Year. Journal of Pediatrics 2020: 223: 20-+.

Sharma A, Xin YM, Chen XG, Sood BG. Early prediction of moderate to severe bronchopulmonary dysplasia in extremely premature infants. Pediatrics and Neonatology 2020: 61(3): 290-299.

Sotodate G, Oyama K, Matsumoto A, Konishi Y, Toya Y, Takashimizu N. Predictive ability of neonatal illness severity scores for early death in extremely premature infants. Journal of Maternal-Fetal & Neonatal Medicine 2020: 1-6.

Baud O, Laughon M, Lehert P. Survival without Bronchopulmonary Dysplasia of Extremely Preterm Infants: A Predictive Model at Birth. Neonatology 2021: 1-9.

Imai K, Nakano-Kobayashi T, Nakamura N, Kajiyama H, Ushida T, Kotani T, Moriyama Y, Nakatochi M, Kobayashi Y, Hayakawa M. Antenatal prediction models for short- and medium-term outcomes in preterm infants. Acta Obstetricia et Gynecologica Scandinavica 2021: 100(6): 1089-1096.

Rysavy MA, Li L, Tyson JE, Jensen EA, Das A, Ambalavanan N, Laughon MM, Greenberg RG, Patel RM, Pedroza C, et al. Should Vitamin A Injections to Prevent Bronchopulmonary Dysplasia or Death Be Reserved for High-Risk Infants?: reanalysis of the NICHD Neonatal Research Network Randomized Trial. Journal of pediatrics 2021.

Rysavy M, Li L, Bell E. Estimating the effect of vitamin a therapy to prevent bronchopulmonary dysplasia or death to very low birth weight infants born two decades after a clinical trial. Paediatric and Perinatal Epidemiology 2021: 35(SUPPL 1): 32-33.

Shim SY, Yun JY, Cho SJ, Kim MH, Park EA. The Prediction of Bronchopulmonary Dysplasia in Very Low Birth Weight Infants through Clinical Indicators within 1 Hour of Delivery. Journal of Korean Medical Science 2021: 36(11): 12.

Sucasas Alonso A, Pertega Diaz S, Saez Soto R, Avila-Alvarez A. [Epidemiology and risk factors for bronchopulmonary dysplasia in prematures infants born at or less than 32 weeks of gestation]. Epidemiologia y factores de riesgo asociados a displasia broncopulmonar en prematuros menores de 32 semanas de edad gestacional 2021.

#### Appendix 5: Characteristics of 53 BPD prediction models included. BMI = Body mass index. PROM = Prolonged rupture of membrane. RDS = Respiratory distress syndrome. PIE = Pulmonary interstitial emphysema. IVH = Intraventricular haemorrhage. FiO_2_ = Fraction of inspired oxygen. PaO_2_ = Arterial partial pressure of oxygen. PaCO_2_ = Arterial partial pressure of carbon dioxide. A-a DO_2_ = Alveolar-arterial partial pressure of oxygen difference. CC = Clinical consensus. NR = Not recorded. D = Day. H = Hour. M = Minutes. EXT = Extubation. DUR = Duration. T = Trend. DESAT = Desaturation. SIG = Significant. HFOV = High frequency oscillation ventilation. CUM = cumulative. EOS = Eosinophil. NUT = Neutrophil.

| **Study** | Cohen 1983 | | Palta 1990 | | Parker 1992 | | Corcoran 1993 | | | | Ryan 1994 | | Gortner 1996 | | | Rozycki 1996 | | | | | | Ryan 1996 | | | | | | Romagnoli 1998 | | | Yoder 1999 | | | Chien 2002 | | Groves 2004 | | | Cunha 2005 | | Kim 2005 | | | Choi 2006 | | | Henderson-Smart 2006 | | | | | Bhering 2007 | | | May 2007 | | | Ambalavanan 2008 | | |
| --- | --- | --- | --- | --- | --- | --- | --- | --- | --- | --- | --- | --- | --- | --- | --- | --- | --- | --- | --- | --- | --- | --- | --- | --- | --- | --- | --- | --- | --- | --- | --- | --- | --- | --- | --- | --- | --- | --- | --- | --- | --- | --- | --- | --- | --- | --- | --- | --- | --- | --- | --- | --- | --- | --- | --- | --- | --- | --- | --- | --- |
|  |  |  |  |  |  |  |  |  |  |  |  |  |  |  |  |  |  |  |  |  |  | BPD 28d | | | BPD 36w | | |  |  |  |  |  |  |  |  |  |  |  |  |  |  |  |  |  |  |  |  |  |  |  |  |  |  |  |  |  |  |  |  |  |
| No predictors considered | CC | | CC | | 10 | | 17 | | | | 12 | | 27 | | | 16 | | | | 19 | | 9 | | | 9 | | | 10 | | | 7 | | | NR | | 6 | | | 16 | | 27 | | | 16 | | | 21 | | | | | 66 | | | NR | | | 26 | | |
| No predictors used | 2 | | 9 | | 7 | | 7 | | | | 4 | | 8 | | | 5 | | | | 3 | | 3 | | | 3 | | | 10 | | | 6 | | | 11 | | 1 | | | 4 | | 8 | | | 3 | | | 3 | | | | | 4 | | | 2 | | | 6 | | |
| Age at prediction | 2d | | 2-3d | | 1d | | 3d | | | | 7d | | 7d | | | 8h | | | | 14d | | 4d | | | 4d | | | 3, 5d | | | 12h, 3d | | | 12h | | 14d | | | 7d | | 4, 7, 10d | | | 7d | | | birth | | | | | 7d | | | 2, 7d | | | 1d | | |
| Gestational age |  | |  | | ✓ | | ✓ | | | | ✓ | | ✓ | | | ✓ | | | |  | |  | | |  | | | ✓ | | | ✓ | | | ✓ | |  | | | ✓ | | ✓ | | | ✓ | | | ✓ | | | | | ✓ | | |  | | |  | | |
| Birthweight | ✓ | | ✓ | | ✓ | | ✓ | | | |  | | ✓ | | |  | | | |  | | ✓ | | | ✓ | | | ✓ | | | ✓ | | |  | |  | | |  | | ✓ | | |  | | | ✓ | | | | |  | | |  | | | ✓ | | |
| Small for gestational age |  | |  | |  | |  | | | |  | |  | | |  | | | |  | |  | | |  | | |  | | |  | | | ✓ | |  | | |  | |  | | |  | | |  | | | | |  | | |  | | |  | | |
| Gender |  | |  | | ✓ | | ✓ | | | |  | | ✓ | | |  | | | |  | |  | | |  | | |  | | |  | | | ✓ | |  | | |  | |  | | |  | | | ✓ | | | | |  | | |  | | | ✓ | | |
| Maternal age |  | |  | |  | |  | | | |  | |  | | |  | | | |  | |  | | |  | | |  | | |  | | |  | |  | | |  | |  | | |  | | |  | | | | |  | | |  | | |  | | |
| Maternal BMI |  | |  | |  | |  | | | |  | |  | | |  | | | |  | |  | | |  | | |  | | |  | | |  | |  | | |  | |  | | |  | | |  | | | | |  | | |  | | |  | | |
| Maternal hypertension |  | |  | |  | |  | | | |  | |  | | |  | | | |  | |  | | |  | | |  | | |  | | |  | |  | | |  | |  | | |  | | |  | | | | |  | | |  | | |  | | |
| Multiple gestation |  | |  | |  | |  | | | |  | |  | | |  | | | |  | |  | | |  | | |  | | |  | | |  | |  | | |  | |  | | |  | | |  | | | | |  | | |  | | |  | | |
| Chorioamnionitis |  | |  | |  | |  | | | |  | |  | | |  | | | |  | |  | | |  | | |  | | |  | | |  | |  | | |  | |  | | |  | | |  | | | | |  | | |  | | |  | | |
| PROM |  | |  | |  | |  | | | |  | |  | | |  | | | |  | |  | | |  | | |  | | |  | | |  | |  | | |  | |  | | |  | | |  | | | | |  | | |  | | |  | | |
| Race |  | |  | | ✓ | |  | | | |  | |  | | |  | | | |  | |  | | |  | | |  | | |  | | |  | |  | | |  | |  | | |  | | |  | | | | |  | | |  | | |  | | |
| Apgar |  | | ✓5m | | ✓5m | |  | | | |  | |  | | | ✓5m | | | |  | |  | | |  | | |  | | |  | | | ✓5m | |  | | |  | | ✓5m | | |  | | |  | | | | |  | | |  | | |  | | |
| Outborn |  | |  | | ✓ | |  | | | |  | |  | | |  | | | |  | |  | | |  | | |  | | |  | | | ✓ | |  | | |  | |  | | |  | | |  | | | | |  | | |  | | | ✓ | | |
| Delivery mode |  | |  | |  | |  | | | |  | | ✓ | | |  | | | |  | |  | | |  | | |  | | |  | | |  | |  | | |  | |  | | |  | | |  | | | | |  | | |  | | |  | | |
| Antenatal steroids |  | |  | |  | |  | | | |  | |  | | |  | | | |  | |  | | |  | | |  | | |  | | |  | |  | | |  | |  | | |  | | |  | | | | |  | | |  | | |  | | |
| Surfactant |  | |  | |  | |  | | | |  | | ✓ | | |  | | | |  | |  | | |  | | |  | | |  | | |  | |  | | |  | |  | | |  | | |  | | | | |  | | |  | | | ✓ | | |
| Mean blood pressure |  | |  | |  | |  | | | |  | |  | | |  | | | |  | |  | | |  | | |  | | |  | | | ✓ | |  | | |  | |  | | |  | | |  | | | | |  | | |  | | |  | | |
| Mean oxygen saturation |  | |  | |  | |  | | | |  | |  | | |  | | | |  | |  | | |  | | |  | | |  | | |  | |  | | |  | |  | | |  | | |  | | | | |  | | |  | | |  | | |
| Heart rate |  | |  | |  | |  | | | |  | |  | | |  | | | |  | |  | | |  | | |  | | |  | | |  | |  | | |  | |  | | |  | | |  | | | | |  | | |  | | |  | | |
| Lowest temperature |  | |  | |  | |  | | | |  | |  | | |  | | | |  | |  | | |  | | |  | | |  | | | ✓ | |  | | |  | |  | | |  | | |  | | | | |  | | |  | | |  | | |
| Urine output |  | |  | |  | |  | | | |  | |  | | |  | | | |  | |  | | |  | | |  | | |  | | | ✓ | |  | | |  | |  | | |  | | |  | | | | |  | | |  | | |  | | |
| Weight loss |  | |  | |  | |  | | | |  | | ✓ | | |  | | | |  | |  | | |  | | |  | | |  | | |  | |  | | |  | |  | | |  | | |  | | | | | ✓ | | |  | | |  | | |
| Fluid intake |  | |  | |  | |  | | | |  | | ✓ | | |  | | | |  | |  | | |  | | |  | | |  | | |  | |  | | | ✓ | |  | | |  | | |  | | | | |  | | |  | | |  | | |
| RDS |  | |  | | ✓ | |  | | | |  | |  | | |  | | | |  | |  | | |  | | | ✓ | | |  | | |  | |  | | |  | |  | | |  | | |  | | | | |  | | |  | | |  | | |
| Pneumonia |  | |  | |  | | ✓ | | | |  | |  | | |  | | | |  | |  | | |  | | |  | | |  | | |  | |  | | |  | |  | | |  | | |  | | | | |  | | |  | | |  | | |
| Pneumothorax |  | | ✓ | |  | |  | | | |  | |  | | |  | | | |  | |  | | |  | | |  | | |  | | |  | |  | | |  | |  | | |  | | |  | | | | |  | | |  | | |  | | |
| Pulmonary haemorrhage |  | | ✓ | |  | |  | | | |  | |  | | |  | | | |  | |  | | |  | | |  | | |  | | |  | |  | | |  | |  | | |  | | |  | | | | |  | | |  | | |  | | |
| PIE |  | | ✓ | |  | |  | | | |  | |  | | |  | | | |  | |  | | |  | | | ✓ | | |  | | |  | |  | | |  | |  | | |  | | |  | | | | |  | | |  | | |  | | |
| Patent ductus arteriosus |  | |  | |  | |  | | | | ✓ | |  | | |  | | | |  | | ✓ | | |  | | | ✓ | | |  | | |  | |  | | | ✓ | |  | | |  | | |  | | | | | ✓ | | |  | | |  | | |
| Sepsis |  | |  | |  | |  | | | |  | |  | | |  | | | | ✓ | |  | | |  | | | ✓ | | |  | | |  | |  | | |  | |  | | |  | | |  | | | | |  | | |  | | |  | | |
| Seizure |  | |  | |  | |  | | | |  | |  | | |  | | | |  | |  | | |  | | |  | | |  | | | ✓ | |  | | |  | |  | | |  | | |  | | | | |  | | |  | | |  | | |
| IVH |  | |  | |  | |  | | | |  | |  | | |  | | | |  | |  | | |  | | | ✓ | | |  | | |  | |  | | |  | |  | | |  | | |  | | | | |  | | |  | | |  | | |
| Congenital anomaly |  | |  | |  | |  | | | |  | |  | | |  | | | |  | |  | | |  | | |  | | |  | | |  | |  | | |  | |  | | |  | | |  | | | | |  | | |  | | |  | | |
| Diuretics |  | |  | |  | |  | | | |  | |  | | |  | | | |  | |  | | |  | | |  | | |  | | |  | |  | | |  | |  | | |  | | |  | | | | |  | | |  | | |  | | |
| Postnatal steroids |  | |  | |  | |  | | | |  | |  | | |  | | | |  | |  | | |  | | |  | | |  | | |  | |  | | |  | |  | | |  | | |  | | | | |  | | |  | | |  | | |
| Centre effect |  | |  | |  | |  | | | |  | |  | | |  | | | |  | |  | | |  | | |  | | |  | | |  | |  | | |  | |  | | |  | | |  | | | | |  | | |  | | |  | | |
| Chest X ray |  | | ✓ | |  | |  | | | |  | |  | | |  | | | |  | |  | | |  | | |  | | |  | | |  | |  | | |  | |  | | |  | | |  | | | | |  | | |  | | |  | | |
| **Ventilator settings** |  | |  | |  | |  | | | |  | |  | | |  | |  | | | |  | | |  | | |  | | |  | | |  | |  | | |  | |  | | |  | | |  | | | | |  | | |  | | |  | | |
| Modality |  | |  | |  | |  | | | |  | |  | | |  | |  | | | |  | | |  | | |  | | |  | | |  | |  | | |  | | ✓ | | |  | | |  | | | | |  | | | ✓ | | |  | | |
| Invasive ventilation | ✓ | |  | |  | | ✓AGE | | | | ✓ | |  | | |  | |  | | | | ✓ | | | ✓ | | | ✓ | | |  | | |  | |  | | |  | |  | | |  | | |  | | | | | ✓>2D | | |  | | |  | | |
| Delivery room intubation |  | |  | |  | |  | | | |  | |  | | |  | |  | | | |  | | |  | | |  | | |  | | |  | |  | | |  | |  | | |  | | |  | | | | |  | | |  | | |  | | |
| Mean F_i_O_2_ | ✓≥0.6 | | ✓MAX | |  | | ✓≥0.6 | | | | ✓MAX | | ✓MAX | | |  | | ✓ | | | |  | | |  | | | ✓ | | | ✓ | | | ✓ | | ✓ | | |  | | ✓ | | | ✓MAX | | |  | | | | |  | | | ✓ | | | ✓ | | |
| Peak inspiratory pressure |  | | ✓ | |  | | ✓>25 | | | |  | |  | | | ✓ | | ✓ | | | |  | | | ✓ | | | ✓ | | | ✓ | | |  | |  | | | ✓ | | ✓ | | |  | | |  | | | | |  | | |  | | |  | | |
| Peak expiratory pressure |  | |  | |  | |  | | | |  | |  | | | ✓ | | ✓ | | | |  | | |  | | |  | | |  | | |  | |  | | |  | |  | | |  | | |  | | | | |  | | |  | | |  | | |
| Mean airway pressure |  | |  | |  | |  | | | |  | |  | | |  | |  | | | |  | | |  | | |  | | | ✓ | | |  | |  | | |  | | ✓ | | | ✓MAX | | |  | | | | |  | | |  | | |  | | |
| Rate |  | |  | |  | |  | | | |  | |  | | | ✓ | | ✓ | | | |  | | |  | | |  | | | ✓ | | |  | |  | | |  | |  | | |  | | |  | | | | |  | | |  | | |  | | |
| **Bloods** |  | |  | |  | |  | | | |  | |  | | |  | |  | | | |  | | |  | | |  | | |  | | |  | |  | | |  | |  | | |  | | |  | | | | |  | | |  | | |  | | |
| pH |  | |  | |  | |  | | | |  | |  | | |  | |  | | | |  | | |  | | |  | | |  | | | ✓ | |  | | |  | |  | | |  | | |  | | | | |  | | |  | | |  | | |
| p_a_O_2_ |  | | ✓ | |  | |  | | | |  | |  | | |  | |  | | | |  | | |  | | |  | | |  | | | ✓ | |  | | |  | | ✓ | | |  | | |  | | | | |  | | |  | | | ✓ | | |
| p_a_CO_2_ |  | |  | |  | |  | | | |  | |  | | | ✓ | | ✓ | | | |  | | |  | | |  | | |  | | |  | |  | | |  | |  | | |  | | |  | | | | |  | | |  | | |  | | |
| A-a DO_2_ |  | |  | |  | |  | | | |  | |  | | |  | |  | | | |  | | |  | | |  | | |  | | |  | |  | | |  | |  | | |  | | |  | | | | |  | | |  | | |  | | |
| Base Excess |  | |  | |  | |  | | | |  | |  | | |  | |  | | | |  | | |  | | |  | | |  | | |  | |  | | |  | |  | | |  | | |  | | | | |  | | |  | | |  | | |
| White blood cell |  | |  | |  | |  | | | |  | |  | | |  | |  | | | |  | | |  | | |  | | |  | | |  | |  | | |  | |  | | |  | | |  | | | | |  | | |  | | |  | | |
| **Study** | Laughon 2011 | | Gottipati 2012 | | | Roth-Kleiner  2012 | | | Gursoy 2014 | | | Yang 2014 | | | Anand 2015 | | Ochab 2016 | | | | | | | Wai 2016 | | | Kim 2017 | | Beltempo 2018 | | | Boghossian 2018 | | | Hunt 2018 | | Sullivan 2018 | | | Fairchild 2019 | | | Sun 2019 | | Valenzuela–Stutman 2019 | | | | | | | | | Dylag 2020 | | | Mistry 2020 | | | Shah 2020 |
|  |  |  |  |  |  |  |  |  |  |  |  |  |  |  |  |  | LR | | | | SVM | | |  |  |  |  |  |  |  |  |  |  |  |  |  |  |  |  |  |  |  |  |  |  |  |  |  |  |  |  |  |  |  |  |  |  |  |  |  |
| No predictors considered | 11 | | 16 | | | NR | | | 31 | | | 1 | | | NR | | 14 | | | | 14 | | | 14 | | | 14 | | 10 | | | 2 | | | 1 | | 23 | | | 13 | | | 2 | | 12 | | | 17 | | 18 | | | 21 | NR | | | 13 | | | NR |
| No predictors used | 6 | | 6 | | | 5 | | | 7 | | | 1 | | | 5 | | 5 | | | | 9 | | | 6 | | | 6 | | 10 | | | 2 | | | 1 | | 23 | | | 5 | | | 1 | | NR (top 5) | | | | | | | | | 1 | | | 11 | | | 5 |
| Age at prediction | 1, 3, 7, 14d | | NR | | | 7d | | | 3d | | | 8-11d | | | birth | | 7d | | | | 7d | | | 14d | | | 7d | | 12h | | | birth | | | 7d | | 7d | | | 7d | | | 3d | | birth | | | 3d | | 7d | | | 14d | 14d | | | 14d | | | 1d |
| Gestational age | ✓ | | ✓ | | | ✓ | | | ✓ | | |  | | | ✓ | | ✓ | | | | ✓ | | |  | | | ✓ | | ✓ | | | ✓ | | |  | | ✓ | | | ✓ | | |  | | ✓ | | |  | |  | | |  |  | | | ✓ | | | ✓ |
| Birthweight | ✓ | |  | | | ✓ | | | ✓ | | |  | | | ✓ | |  | | | |  | | |  | | | ✓ | | ✓ | | |  | | |  | | ✓ | | | ✓ | | |  | | ✓ | | | ✓ | | ✓ | | |  |  | | | ✓ | | |  |
| Small for gestational age |  | |  | | |  | | |  | | |  | | |  | |  | | | |  | | |  | | |  | |  | | | ✓ | | |  | |  | | |  | | |  | |  | | |  | |  | | | ✓ |  | | |  | | |  |
| Gender | ✓ | |  | | |  | | | ✓ | | |  | | | ✓ | |  | | | |  | | |  | | |  | | ✓ | | |  | | |  | | ✓ | | |  | | |  | | ✓ | | | ✓ | | ✓ | | | ✓ |  | | | ✓ | | | ✓ |
| Maternal age |  | |  | | |  | | |  | | |  | | | ✓ | |  | | | |  | | |  | | |  | |  | | |  | | |  | |  | | |  | | |  | |  | | |  | |  | | |  |  | | |  | | |  |
| Maternal BMI |  | |  | | |  | | |  | | |  | | | ✓ | |  | | | |  | | |  | | |  | |  | | |  | | |  | |  | | |  | | |  | |  | | |  | |  | | |  |  | | |  | | |  |
| Maternal hypertension |  | |  | | |  | | |  | | |  | | |  | |  | | | |  | | |  | | |  | |  | | |  | | |  | |  | | |  | | |  | |  | | | ✓ | |  | | |  |  | | |  | | | ✓ |
| Multiple gestation |  | |  | | |  | | |  | | |  | | |  | |  | | | |  | | |  | | |  | |  | | |  | | |  | |  | | |  | | |  | |  | | |  | |  | | |  |  | | |  | | |  |
| Chorioamnionitis |  | |  | | |  | | |  | | |  | | |  | |  | | | |  | | |  | | |  | |  | | |  | | |  | |  | | |  | | |  | |  | | |  | |  | | |  |  | | |  | | |  |
| PROM |  | |  | | |  | | |  | | |  | | |  | |  | | | |  | | |  | | |  | |  | | |  | | |  | |  | | |  | | |  | |  | | |  | |  | | |  |  | | |  | | |  |
| Race | ✓ | |  | | |  | | |  | | |  | | |  | |  | | | |  | | |  | | |  | |  | | |  | | |  | |  | | |  | | |  | |  | | |  | |  | | |  |  | | |  | | |  |
| Apgar |  | |  | | |  | | |  | | |  | | |  | |  | | | |  | | |  | | |  | |  | | |  | | |  | | ✓1/5m | | |  | | |  | | ✓1m | | |  | |  | | |  |  | | |  | | |  |
| Outborn |  | |  | | |  | | |  | | |  | | |  | |  | | | |  | | |  | | |  | |  | | |  | | |  | |  | | |  | | |  | |  | | |  | |  | | |  |  | | |  | | |  |
| Delivery mode |  | |  | | |  | | |  | | |  | | |  | |  | | | |  | | |  | | |  | |  | | |  | | |  | |  | | |  | | |  | |  | | |  | |  | | |  |  | | |  | | |  |
| Antenatal steroids |  | |  | | |  | | |  | | |  | | |  | |  | | | |  | | |  | | |  | |  | | |  | | |  | | ✓ | | |  | | |  | |  | | |  | |  | | |  |  | | | ✓ | | |  |
| Surfactant |  | | ✓ | | |  | | |  | | |  | | |  | |  | | | |  | | |  | | |  | |  | | |  | | |  | |  | | |  | | |  | |  | | |  | |  | | |  |  | | | ✓ | | |  |
| Mean blood pressure |  | |  | | |  | | | ✓ | | |  | | |  | | ✓&T | | | | ✓&T | | |  | | |  | | ✓ | | |  | | |  | |  | | |  | | |  | |  | | |  | |  | | |  |  | | |  | | |  |
| Mean oxygen saturation |  | |  | | |  | | |  | | |  | | |  | | ✓85 | | | | ✓&T/85/95 | | |  | | |  | |  | | |  | | |  | | ✓T | | | ✓DESAT | | |  | |  | | |  | |  | | |  |  | | |  | | |  |
| Heart rate |  | |  | | |  | | |  | | |  | | |  | |  | | | |  | | |  | | |  | |  | | |  | | |  | | ✓T | | |  | | |  | |  | | |  | |  | | |  |  | | |  | | |  |
| Lowest temperature |  | |  | | |  | | |  | | |  | | |  | |  | | | |  | | |  | | |  | | ✓ | | |  | | |  | |  | | |  | | |  | |  | | |  | |  | | |  |  | | |  | | |  |
| Urine output |  | |  | | |  | | |  | | |  | | |  | |  | | | |  | | |  | | |  | | ✓ | | |  | | |  | |  | | |  | | |  | |  | | |  | |  | | |  |  | | |  | | |  |
| Weight loss |  | |  | | |  | | |  | | |  | | |  | |  | | | |  | | |  | | |  | |  | | |  | | |  | |  | | |  | | |  | |  | | |  | |  | | |  |  | | |  | | |  |
| Fluid intake |  | |  | | |  | | |  | | |  | | |  | |  | | | |  | | |  | | |  | |  | | |  | | |  | |  | | |  | | |  | |  | | |  | |  | | |  |  | | |  | | |  |
| RDS |  | |  | | |  | | | ✓ | | |  | | |  | |  | | | |  | | |  | | | ✓ | |  | | |  | | |  | |  | | |  | | |  | |  | | | ✓ | | ✓ | | | ✓ |  | | |  | | |  |
| Pneumonia |  | |  | | |  | | |  | | |  | | |  | |  | | | |  | | |  | | | ✓ | |  | | |  | | |  | |  | | |  | | |  | |  | | |  | |  | | |  |  | | |  | | |  |
| Pneumothorax |  | |  | | |  | | |  | | |  | | |  | |  | | | |  | | |  | | |  | |  | | |  | | |  | |  | | |  | | |  | |  | | |  | |  | | |  |  | | |  | | |  |
| Pulmonary haemorrhage |  | |  | | |  | | |  | | |  | | |  | |  | | | |  | | |  | | |  | |  | | |  | | |  | |  | | |  | | |  | |  | | |  | |  | | |  |  | | |  | | |  |
| PIE |  | |  | | |  | | |  | | |  | | |  | |  | | | |  | | |  | | |  | |  | | |  | | |  | |  | | |  | | |  | |  | | |  | |  | | |  |  | | |  | | |  |
| Patent ductus arteriosus |  | | ✓ | | |  | | | ✓SIG | | |  | | |  | | ✓ | | | | ✓ | | |  | | | ✓SIG | |  | | |  | | |  | |  | | |  | | |  | |  | | |  | | ✓ | | |  |  | | | ✓SIG | | |  |
| Sepsis |  | | ✓ | | | ✓ | | |  | | |  | | |  | |  | | | |  | | |  | | |  | |  | | |  | | |  | |  | | |  | | |  | |  | | |  | |  | | |  |  | | | ✓ | | |  |
| Seizure |  | |  | | |  | | |  | | |  | | |  | |  | | | |  | | |  | | |  | | ✓ | | |  | | |  | |  | | |  | | |  | |  | | |  | |  | | |  |  | | |  | | |  |
| IVH |  | |  | | |  | | | ✓ | | |  | | |  | |  | | | |  | | |  | | |  | |  | | |  | | |  | |  | | |  | | |  | |  | | |  | |  | | |  |  | | |  | | |  |
| Congenital anomaly |  | |  | | |  | | |  | | |  | | |  | |  | | | |  | | |  | | |  | |  | | |  | | |  | |  | | |  | | |  | |  | | |  | |  | | |  |  | | |  | | |  |
| Diuretics |  | |  | | |  | | |  | | |  | | |  | |  | | | |  | | |  | | |  | |  | | |  | | |  | |  | | |  | | |  | |  | | |  | |  | | |  |  | | | ✓ | | |  |
| Postnatal steroids |  | |  | | |  | | |  | | |  | | |  | |  | | | |  | | |  | | |  | |  | | |  | | |  | |  | | |  | | |  | |  | | |  | |  | | |  |  | | | ✓ | | |  |
| Centre effect |  | |  | | |  | | |  | | |  | | |  | |  | | | |  | | |  | | |  | |  | | |  | | |  | |  | | |  | | |  | |  | | |  | |  | | |  |  | | |  | | |  |
| Chest X ray |  | |  | | |  | | |  | | |  | | |  | |  | | | |  | | |  | | |  | |  | | |  | | |  | |  | | |  | | |  | |  | | |  | |  | | |  |  | | |  | | |  |
| **Ventilator settings** |  | |  | | |  | | |  | | |  | | |  | |  | | | |  | | |  | | |  | |  | | |  | | |  | |  | | |  | | |  | |  | | |  | |  | | |  |  | | |  | | |  |
| Modality | ✓ | | ✓EXT | | |  | | |  | | |  | | |  | |  | | | |  | | |  | | |  | |  | | |  | | |  | |  | | | ✓ | | |  | |  | | |  | |  | | |  |  | | |  | | |  |
| Invasive ventilation |  | | ✓EXT | | | ✓DUR | | |  | | |  | | |  | |  | | | |  | | |  | | | ✓ | |  | | |  | | | ✓ | |  | | | ✓DUR | | |  | |  | | | ✓ | | ✓ | | | ✓7D |  | | |  | | |  |
| Delivery room intubation |  | |  | | | ✓ | | |  | | |  | | |  | |  | | | |  | | |  | | |  | |  | | |  | | |  | |  | | |  | | |  | | ✓ | | |  | |  | | |  |  | | |  | | |  |
| Mean F_i_O_2_ | ✓ | |  | | |  | | |  | | |  | | |  | |  | | | |  | | | ✓CUM | | |  | | ✓ | | |  | | |  | |  | | | ✓ | | |  | |  | | |  | |  | | | ✓14D | ✓CUM | | | ✓ | | | ✓ |
| Peak inspiratory pressure |  | |  | | |  | | |  | | |  | | |  | |  | | | |  | | |  | | |  | |  | | |  | | |  | |  | | |  | | |  | |  | | |  | |  | | |  |  | | |  | | |  |
| Peak expiratory pressure |  | |  | | |  | | |  | | |  | | |  | |  | | | |  | | |  | | |  | |  | | |  | | |  | |  | | |  | | |  | |  | | |  | |  | | |  |  | | |  | | |  |
| Mean airway pressure |  | |  | | |  | | |  | | |  | | |  | |  | | | |  | | | ✓CUM | | |  | |  | | |  | | |  | |  | | |  | | |  | |  | | |  | |  | | |  |  | | | ✓ | | | ✓ |
| Rate |  | |  | | |  | | |  | | |  | | |  | |  | | | |  | | |  | | |  | |  | | |  | | |  | |  | | |  | | |  | |  | | |  | |  | | |  |  | | |  | | |  |
| **Bloods** |  | |  | | |  | | |  | | |  | | |  | |  | | | |  | | |  | | |  | |  | | |  | | |  | |  | | |  | | |  | |  | | |  | |  | | |  |  | | |  | | |  |
| pH |  | |  | | |  | | |  | | |  | | |  | |  | | | |  | | |  | | |  | | ✓ | | |  | | |  | |  | | |  | | |  | |  | | |  | |  | | |  |  | | |  | | |  |
| p_a_O_2_ |  | |  | | |  | | |  | | |  | | |  | |  | | | |  | | |  | | |  | | ✓ | | |  | | |  | |  | | |  | | |  | |  | | |  | |  | | |  |  | | |  | | |  |
| p_a_CO_2_ |  | |  | | |  | | |  | | |  | | |  | |  | | | |  | | |  | | |  | |  | | |  | | |  | |  | | |  | | |  | |  | | |  | |  | | |  |  | | |  | | |  |
| A-a DO_2_ |  | |  | | |  | | |  | | |  | | |  | |  | | | | ✓ | | |  | | |  | |  | | |  | | |  | |  | | |  | | |  | |  | | |  | |  | | |  |  | | |  | | |  |
| Base Excess |  | |  | | |  | | |  | | |  | | |  | |  | | | |  | | |  | | |  | |  | | |  | | |  | |  | | |  | | |  | |  | | |  | |  | | |  |  | | |  | | |  |
| White blood cell |  | |  | | |  | | |  | | | ✓EOS | | |  | |  | | | |  | | |  | | |  | |  | | |  | | |  | |  | | |  | | | ✓NUT | |  | | |  | |  | | |  |  | | |  | | |  |
| **Study** | | Sharma 2020 | | Vaid 2020 | | | | Baud 2021 | | Shim 2021 | | | | | | | | | Ushida 2021 | | | | CRIB I | | | Sinkin 1990 | | | | Respiratory Severity Score | | | Berlin Admission Score | | | | | SNAP II | | | | SNAPPE II | | | | CRIB II | | | HRC index | | Srisuparp 2003 | | | | | SpO_2_ / P_i_O_2_ shift | | | **Total**  **(n (%))** | |
|  |  |  |  |  |  |  |  |  |  | BPD 28d | | | | BPD 36w | | | | |  |  |  |  |  |  |  |  |  |  |  |  |  |  |  |  |  |  |  |  |  |  |  |  |  |  |  |  |  |  |  |  |  |  |  |  |  |  |  |  |  |  |
| No predictors considered | | 9 | | 60 | | | | 13 | | 13 | | | | 13 | | | | | 12 | | | | NA | | | NA | | | | NA | | | NA | | | | | NA | | | | NA | | | | NA | | | NA | | NA | | | | | NA | | |  |  |
| No predictors used | | 2 | | 3 | | | | 6 | | 5 | | | | 7 | | | | | 8 | | | | 6 | | | 4 | | | | 2 | | | 5 | | | | | 7 | | | | 9 | | | | 5 | | | 3 | | 3 | | | | | 2 | | |  |  |
| Age at prediction | | 14d | | 1d | | | | 1d | | 1h | | | | 1h | | | | | birth | | | | 12h | | | 12h | | | | Variable | | | Admission | | | | | 12h | | | | 12h | | | | 1h | | | 1, 7d | | 1d | | | | | 7d | | |  |  |
| Gestational age | |  | |  | | | | ✓ | | ✓ | | | | ✓ | | | | | ✓ | | | | ✓ | | | ✓ | | | |  | | |  | | | | |  | | | |  | | | | ✓ | | |  | |  | | | | |  | | | 37 (69) | |
| Birthweight | |  | |  | | | | ✓ | | ✓ | | | | ✓ | | | | | ✓ | | | | ✓ | | | ✓ | | | |  | | | ✓ | | | | |  | | | | ✓ | | | | ✓ | | |  | |  | | | | |  | | | 33 (61) | |
| Small for gestational age | |  | |  | | | |  | |  | | | |  | | | | |  | | | |  | | |  | | | |  | | |  | | | | |  | | | | ✓ | | | |  | | |  | |  | | | | |  | | | 4 (7) | |
| Gender | |  | |  | | | | ✓ | | ✓ | | | | ✓ | | | | | ✓ | | | |  | | |  | | | |  | | |  | | | | |  | | | |  | | | | ✓ | | |  | |  | | | | |  | | | 22 (41) | |
| Maternal age | |  | |  | | | |  | |  | | | |  | | | | |  | | | |  | | |  | | | |  | | |  | | | | |  | | | |  | | | |  | | |  | |  | | | | |  | | | 1 (2) | |
| Maternal BMI | |  | |  | | | |  | |  | | | |  | | | | |  | | | |  | | |  | | | |  | | |  | | | | |  | | | |  | | | |  | | |  | |  | | | | |  | | | 1 (2) | |
| Maternal hypertension | |  | |  | | | |  | |  | | | | ✓ | | | | |  | | | |  | | |  | | | |  | | |  | | | | |  | | | |  | | | |  | | |  | |  | | | | |  | | | 3 (6) | |
| Multiple gestation | |  | |  | | | | ✓ | |  | | | |  | | | | | ✓ | | | |  | | |  | | | |  | | |  | | | | |  | | | |  | | | |  | | |  | |  | | | | |  | | | 2 (4) | |
| Chorioamnionitis | |  | |  | | | |  | |  | | | |  | | | | | ✓ | | | |  | | |  | | | |  | | |  | | | | |  | | | |  | | | |  | | |  | |  | | | | |  | | | 1 (2) | |
| PROM | |  | |  | | | |  | |  | | | |  | | | | | ✓ | | | |  | | |  | | | |  | | |  | | | | |  | | | |  | | | |  | | |  | |  | | | | |  | | | 1 (2) | |
| Race | |  | |  | | | |  | |  | | | |  | | | | |  | | | |  | | |  | | | |  | | |  | | | | |  | | | |  | | | |  | | |  | |  | | | | |  | | | 2 (4) | |
| Apgar | |  | |  | | | |  | | ✓5m | | | | ✓5m | | | | |  | | | |  | | | ✓5m | | | |  | | | ✓5m | | | | |  | | | | ✓5m | | | |  | | |  | |  | | | | |  | | | 12 (22) | |
| Outborn | |  | |  | | | |  | |  | | | |  | | | | |  | | | |  | | |  | | | |  | | |  | | | | |  | | | |  | | | |  | | |  | |  | | | | |  | | | 3 (6) | |
| Delivery mode | |  | |  | | | |  | |  | | | |  | | | | | ✓ | | | |  | | |  | | | |  | | |  | | | | |  | | | |  | | | |  | | |  | |  | | | | |  | | | 2 (4) | |
| Antenatal steroids | |  | |  | | | |  | |  | | | |  | | | | | ✓ | | | |  | | |  | | | |  | | |  | | | | |  | | | |  | | | |  | | |  | |  | | | | |  | | | 3 (6) | |
| Surfactant | |  | | ✓ | | | |  | | ✓ | | | |  | | | | |  | | | |  | | |  | | | |  | | |  | | | | |  | | | |  | | | |  | | |  | |  | | | | |  | | | 6 (11) | |
| Mean blood pressure | |  | |  | | | |  | |  | | | |  | | | | |  | | | |  | | |  | | | |  | | |  | | | | | ✓ | | | | ✓ | | | |  | | |  | |  | | | | |  | | | 5 (9) | |
| Mean oxygen saturation | |  | |  | | | |  | |  | | | |  | | | | |  | | | |  | | |  | | | |  | | |  | | | | |  | | | |  | | | |  | | |  | |  | | | | | ✓ | | | 5 (9) | |
| Heart rate | |  | |  | | | |  | |  | | | |  | | | | |  | | | |  | | |  | | | |  | | |  | | | | |  | | | |  | | | |  | | | ✓T | |  | | | | |  | | | 2 (4) | |
| Lowest temperature | |  | |  | | | |  | |  | | | | ✓ | | | | |  | | | |  | | |  | | | |  | | |  | | | | | ✓ | | | | ✓ | | | | ✓ | | |  | |  | | | | |  | | | 6 (11) | |
| Urine output | |  | |  | | | |  | |  | | | |  | | | | |  | | | |  | | |  | | | |  | | |  | | | | | ✓ | | | | ✓ | | | |  | | |  | |  | | | | |  | | | 4 (7) | |
| Weight loss | |  | |  | | | |  | |  | | | |  | | | | |  | | | |  | | |  | | | |  | | |  | | | | |  | | | |  | | | |  | | |  | |  | | | | |  | | | 2 (4) | |
| Fluid intake | |  | |  | | | |  | |  | | | |  | | | | |  | | | |  | | |  | | | |  | | |  | | | | |  | | | |  | | | |  | | |  | |  | | | | |  | | | 2 (4) | |
| RDS | |  | |  | | | |  | |  | | | | ✓ | | | | |  | | | |  | | |  | | | |  | | | ✓ | | | | |  | | | |  | | | |  | | |  | |  | | | | |  | | | 9 (17) | |
| Pneumonia | |  | |  | | | |  | |  | | | |  | | | | |  | | | |  | | |  | | | |  | | |  | | | | |  | | | |  | | | |  | | |  | |  | | | | |  | | | 2 (4) | |
| Pneumothorax | |  | |  | | | |  | |  | | | |  | | | | |  | | | |  | | |  | | | |  | | |  | | | | |  | | | |  | | | |  | | |  | |  | | | | |  | | | 1 (2) | |
| Pulmonary haemorrhage | |  | |  | | | |  | |  | | | |  | | | | |  | | | |  | | |  | | | |  | | |  | | | | |  | | | |  | | | |  | | |  | |  | | | | |  | | | 1 (2) | |
| PIE | |  | |  | | | |  | |  | | | |  | | | | |  | | | |  | | |  | | | |  | | |  | | | | |  | | | |  | | | |  | | |  | |  | | | | |  | | | 2 (4) | |
| Patent ductus arteriosus | |  | |  | | | |  | |  | | | |  | | | | |  | | | |  | | |  | | | |  | | |  | | | | |  | | | |  | | | |  | | |  | |  | | | | |  | | | 13 (24) | |
| Sepsis | |  | |  | | | |  | |  | | | |  | | | | |  | | | |  | | |  | | | |  | | |  | | | | |  | | | |  | | | |  | | |  | |  | | | | |  | | | 5 (9) | |
| Seizure | |  | |  | | | |  | |  | | | |  | | | | |  | | | |  | | |  | | | |  | | |  | | | | | ✓ | | | | ✓ | | | |  | | |  | |  | | | | |  | | | 4 (7) | |
| IVH | |  | |  | | | |  | |  | | | |  | | | | |  | | | |  | | |  | | | |  | | |  | | | | |  | | | |  | | | |  | | |  | |  | | | | |  | | | 2 (4) | |
| Congenital malformation | |  | |  | | | |  | |  | | | |  | | | | |  | | | | ✓ | | |  | | | |  | | |  | | | | |  | | | |  | | | |  | | |  | |  | | | | |  | | | 1 (2) | |
| Diuretics | |  | |  | | | |  | |  | | | |  | | | | |  | | | |  | | |  | | | |  | | |  | | | | |  | | | |  | | | |  | | |  | |  | | | | |  | | | 1 (2) | |
| Postnatal steroids | |  | | ✓ | | | |  | |  | | | |  | | | | |  | | | |  | | |  | | | |  | | |  | | | | |  | | | |  | | | |  | | |  | |  | | | | |  | | | 2 (4) | |
| Centre effect | |  | |  | | | | ✓ | |  | | | |  | | | | |  | | | |  | | |  | | | |  | | |  | | | | |  | | | |  | | | |  | | |  | |  | | | | |  | | | 1 (2) | |
| Chest X ray | |  | |  | | | |  | |  | | | |  | | | | |  | | | |  | | |  | | | |  | | |  | | | | |  | | | |  | | | |  | | |  | |  | | | | |  | | | 1 (2) | |
| **Ventilator settings** | |  | |  | | | |  | |  | | | |  | | | | |  | | | |  | | |  | | | |  | | |  | | | | |  | | | |  | | | |  | | |  | |  | | | | |  | | |  | |
| Modality | |  | |  | | | | ✓ | |  | | | |  | | | | |  | | | |  | | |  | | | |  | | |  | | | | |  | | | |  | | | |  | | |  | |  | | | | |  | | | 6 (11) | |
| Invasive ventilation | | ✓DUR | |  | | | |  | |  | | | |  | | | | |  | | | |  | | |  | | | |  | | | ✓ | | | | |  | | | |  | | | |  | | |  | |  | | | | |  | | | 17 (33) | |
| Delivery room intubation | |  | | ✓HFOV | | | |  | |  | | | |  | | | | |  | | | |  | | |  | | | |  | | |  | | | | |  | | | |  | | | |  | | |  | |  | | | | |  | | | 3 (6) | |
| Mean F_i_O_2_ | | ✓DUR | |  | | | |  | |  | | | |  | | | | |  | | | | ✓MIN/MAX | | |  | | | | ✓ | | |  | | | | | ✓ | | | | ✓ | | | |  | | |  | |  | | | | | ✓ | | | 28 (52) | |
| Peak inspiratory pressure | |  | |  | | | |  | |  | | | |  | | | | |  | | | |  | | | ✓ | | | |  | | |  | | | | |  | | | |  | | | |  | | |  | |  | | | | |  | | | 9 (17) | |
| Peak expiratory pressure | |  | |  | | | |  | |  | | | |  | | | | |  | | | |  | | |  | | | |  | | |  | | | | |  | | | |  | | | |  | | |  | |  | | | | |  | | | 2 (4) | |
| Mean airway pressure | |  | |  | | | |  | |  | | | |  | | | | |  | | | |  | | |  | | | | ✓ | | |  | | | | |  | | | |  | | | |  | | |  | | ✓ | | | | |  | | | 8 (15) | |
| Rate | |  | |  | | | |  | |  | | | |  | | | | |  | | | |  | | |  | | | |  | | |  | | | | |  | | | |  | | | |  | | |  | |  | | | | |  | | | 3 (6) | |
| **Bloods** | |  | |  | | | |  | |  | | | |  | | | | |  | | | |  | | |  | | | |  | | |  | | | | |  | | | |  | | | |  | | |  | |  | | | | |  | | |  | |
| pH | |  | |  | | | |  | |  | | | |  | | | | |  | | | |  | | |  | | | |  | | |  | | | | | ✓ | | | | ✓ | | | |  | | |  | |  | | | | |  | | | 4 (7) | |
| p_a_O_2_ | |  | |  | | | |  | |  | | | |  | | | | |  | | | |  | | |  | | | |  | | |  | | | | | ✓ | | | | ✓ | | | |  | | |  | | ✓ | | | | |  | | | 8 (15) | |
| p_a_CO_2_ | |  | |  | | | |  | |  | | | |  | | | | |  | | | |  | | |  | | | |  | | |  | | | | |  | | | |  | | | |  | | |  | |  | | | | |  | | | 2 (4) | |
| A-a DO_2_ | |  | |  | | | |  | |  | | | |  | | | | |  | | | |  | | |  | | | |  | | |  | | | | |  | | | |  | | | |  | | |  | | ✓ | | | | |  | | | 2 (4) | |
| Base Excess | |  | |  | | | |  | |  | | | |  | | | | |  | | | | ✓ | | |  | | | |  | | | ✓ | | | | |  | | | |  | | | | ✓ | | |  | |  | | | | |  | | | 3 (6) | |
| White blood cell | |  | |  | | | |  | |  | | | |  | | | | |  | | | |  | | |  | | | |  | | |  | | | | |  | | | |  | | | |  | | |  | |  | | | | |  | | | 2 (4) | |

#### Appendix 6: Risk of bias assessments for included studies based on the PROBAST tool^13^

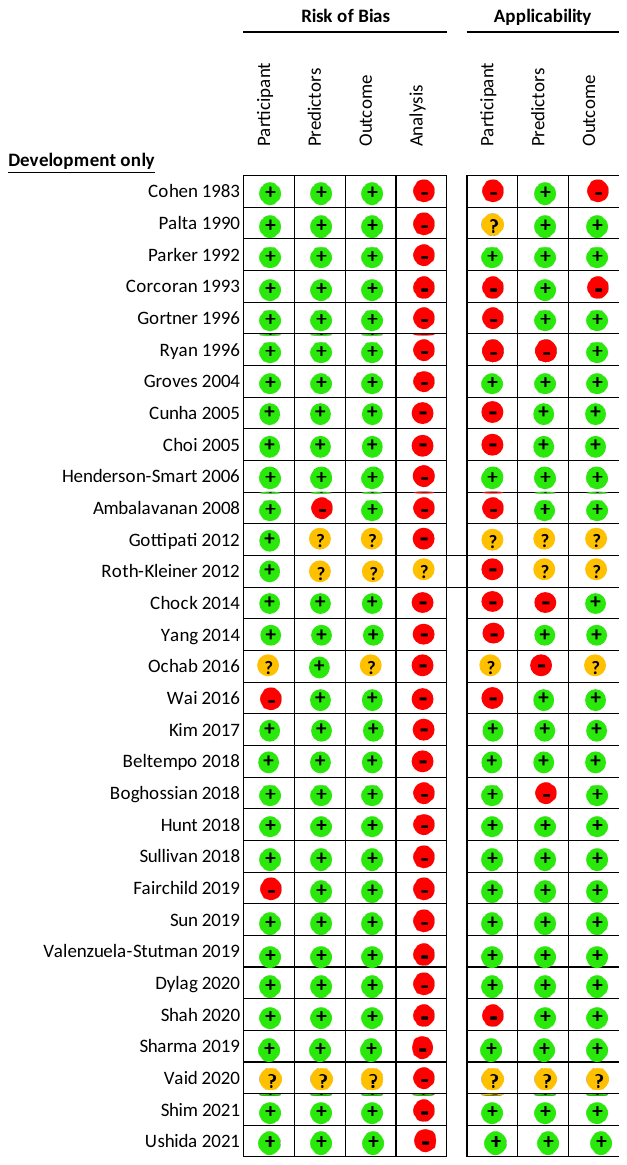

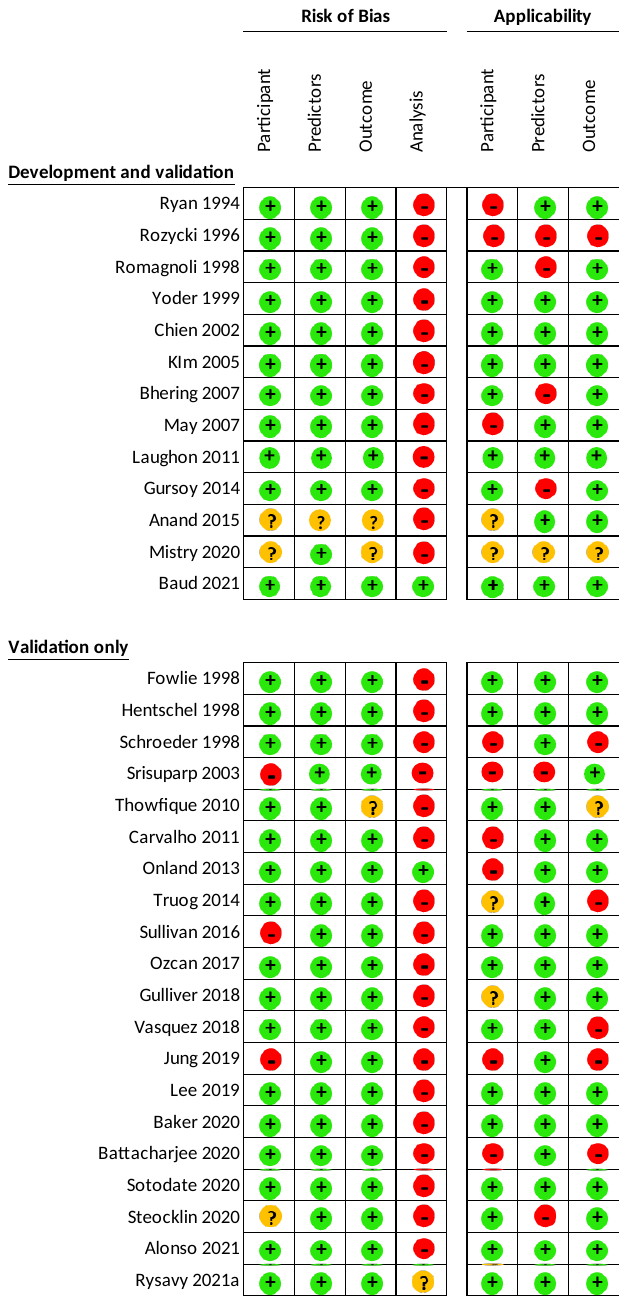

#### Appendix 7: Forest plot of C-statistics in external validation studies of included prediction models for (A) bronchopulmonary dysplasia (BPD) outcome as well as (B) composite outcome of BPD and death.

^1^ BPD was defined as respiratory support requirement at 28 days of age rather than 36 weeks of corrected gestational age.

##### (A) Bronchopulmonary dysplasia

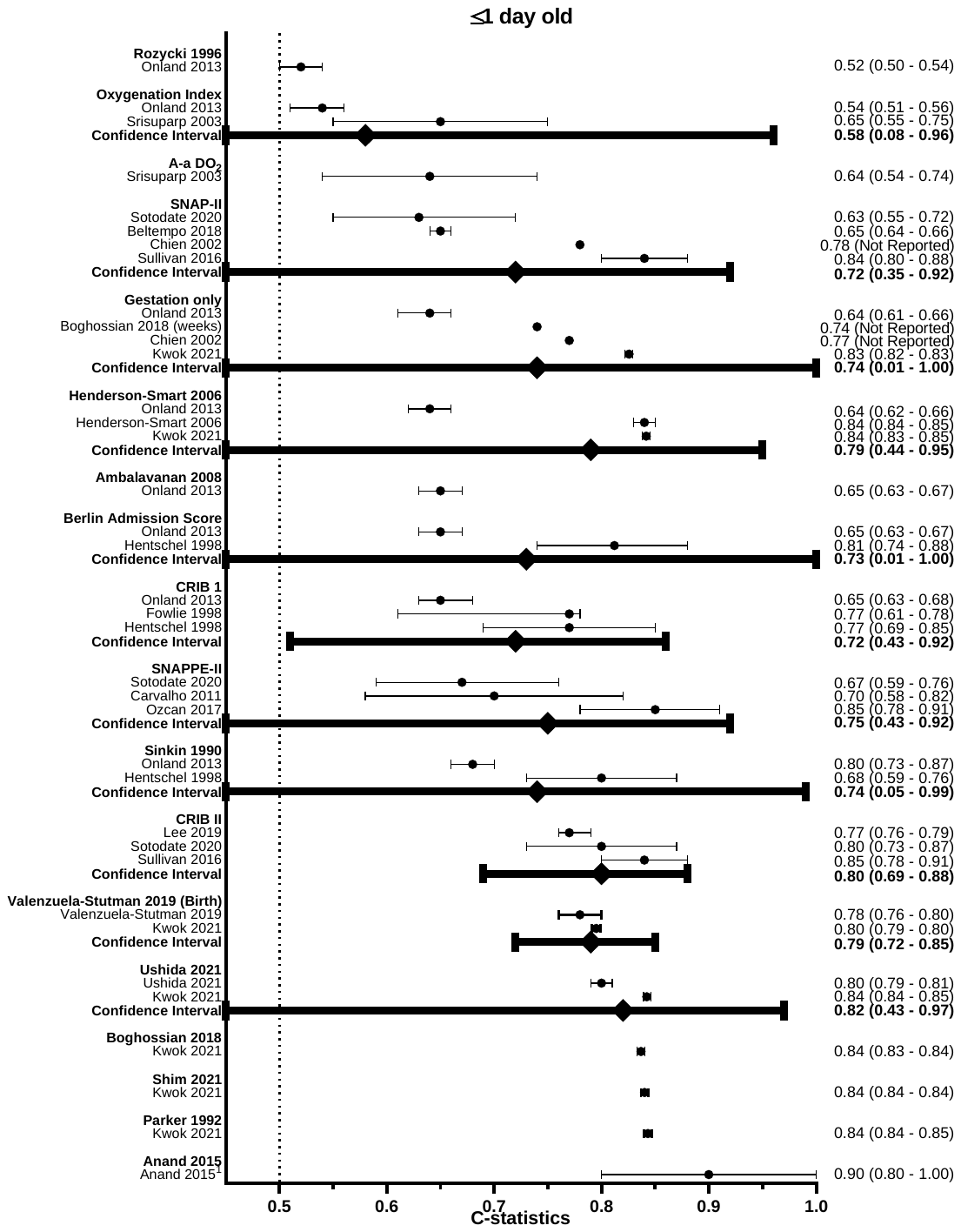

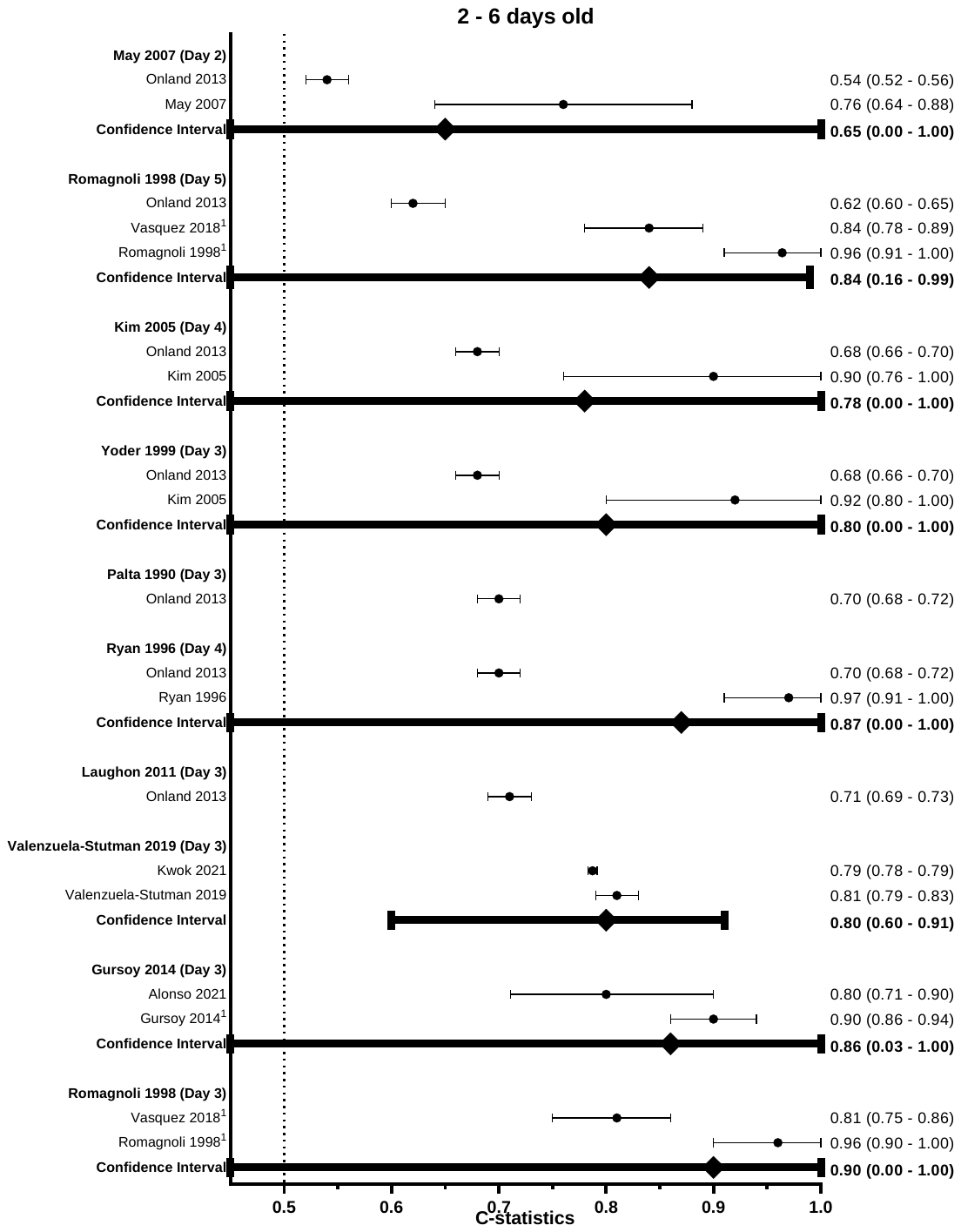

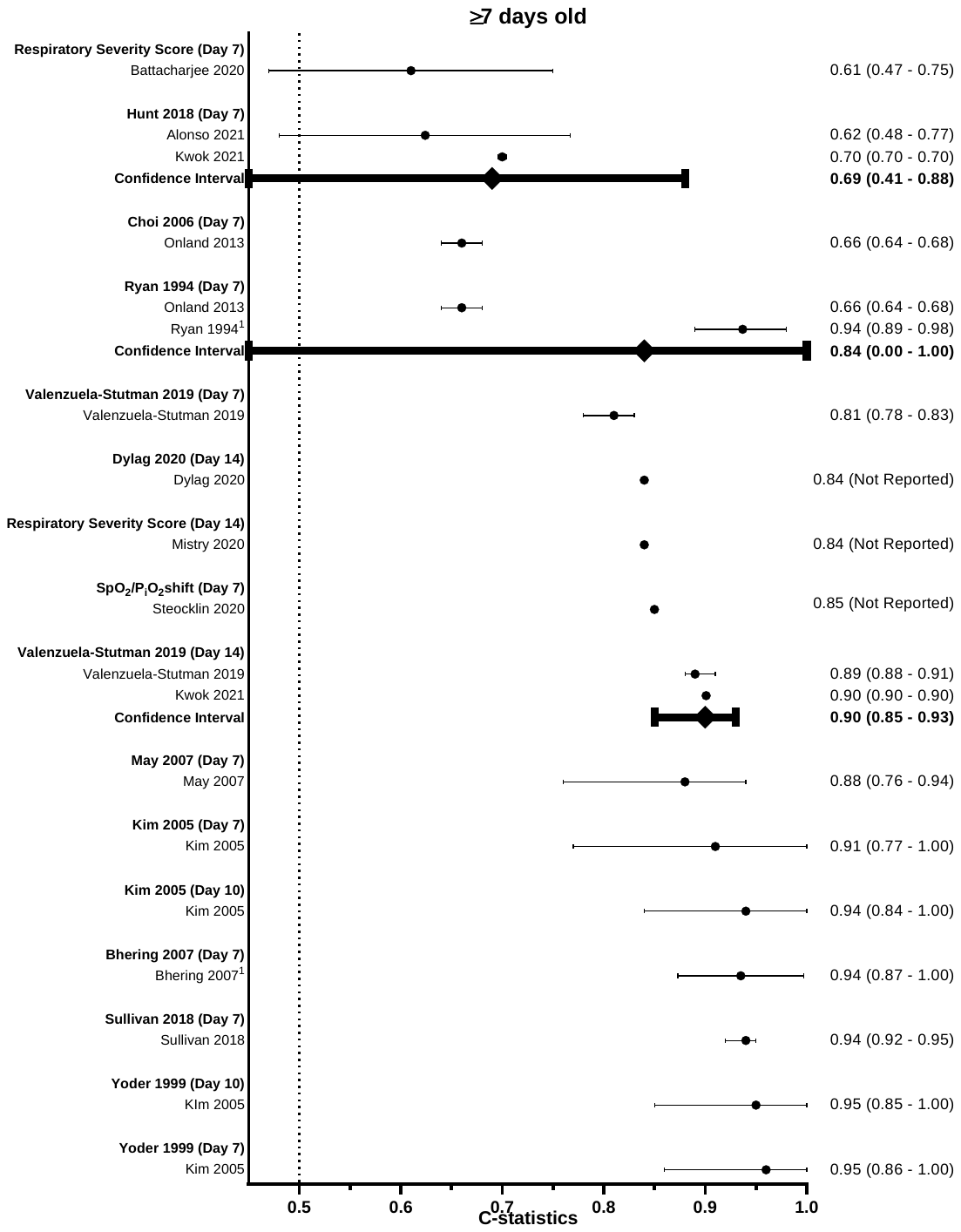

##### (B) Composite outcome of bronchopulmonary dysplasia and death

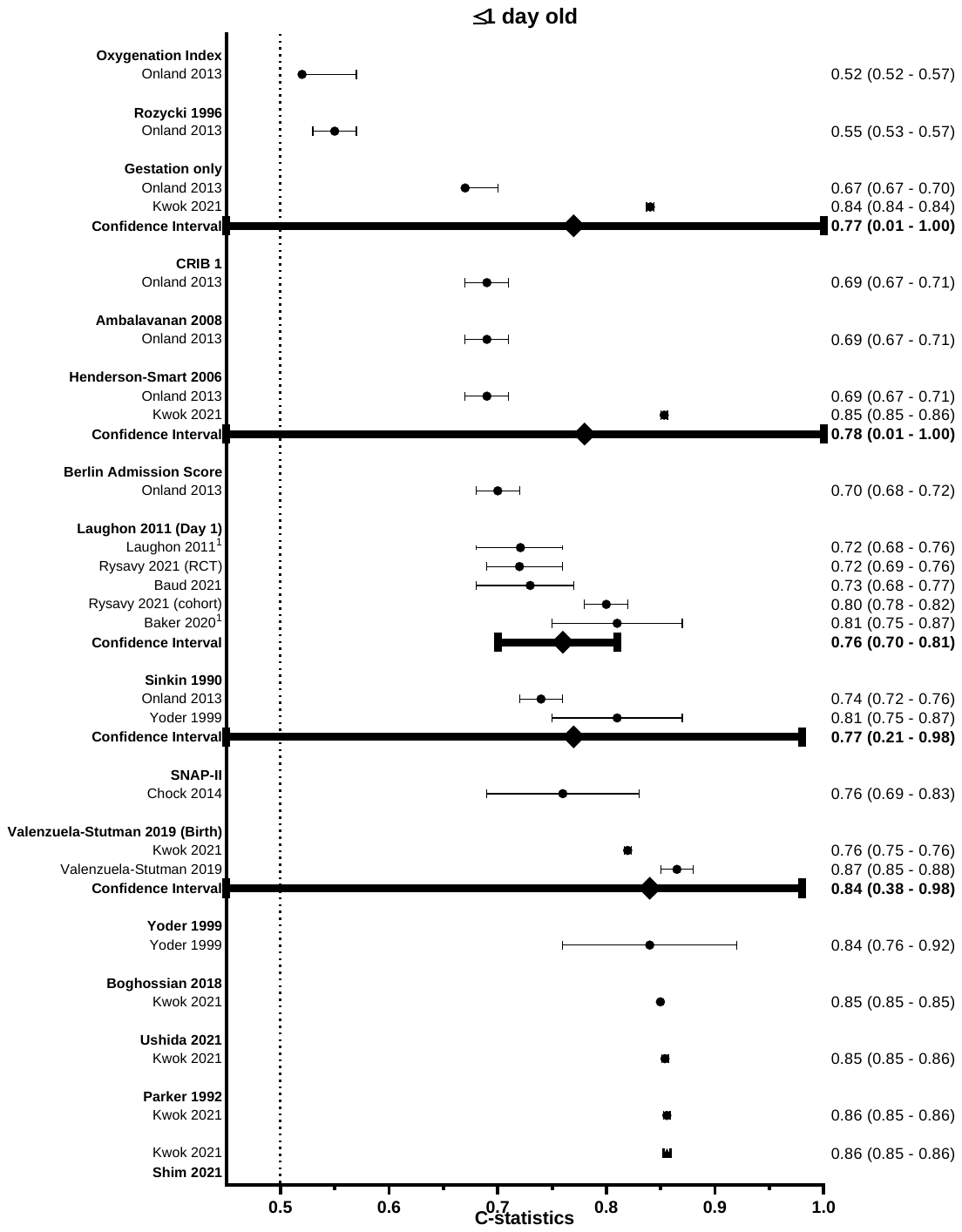

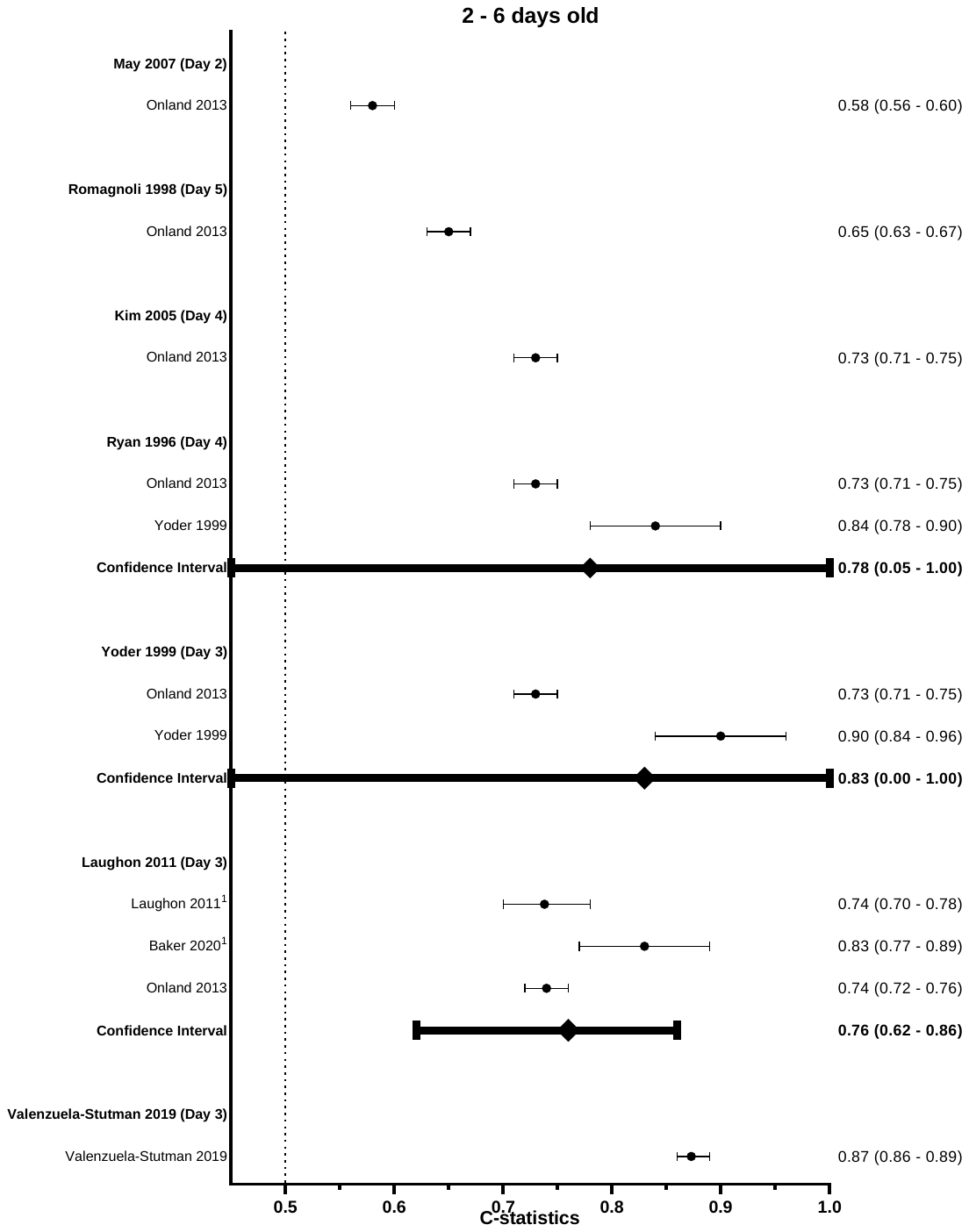

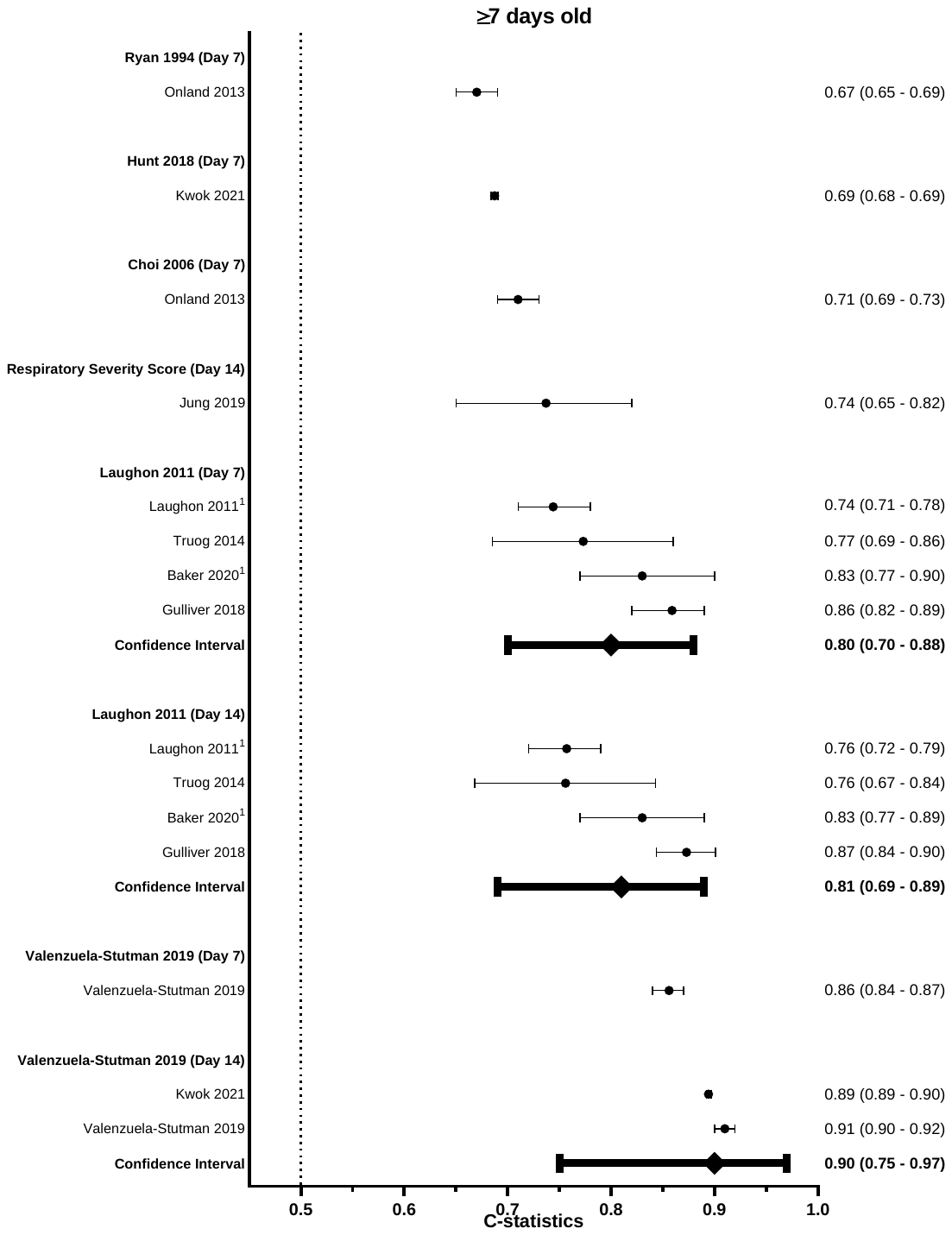

#### Appendix 8: Forest plot of observed : expected ratio (O:E ratio) in external validation studies of included prediction models for (A) bronchopulmonary dysplasia (BPD) outcome as well as (B) composite outcome of BPD and death

^1^ Observed and expected risk were obtained from the observed incidence table stratified by estimated risk group.

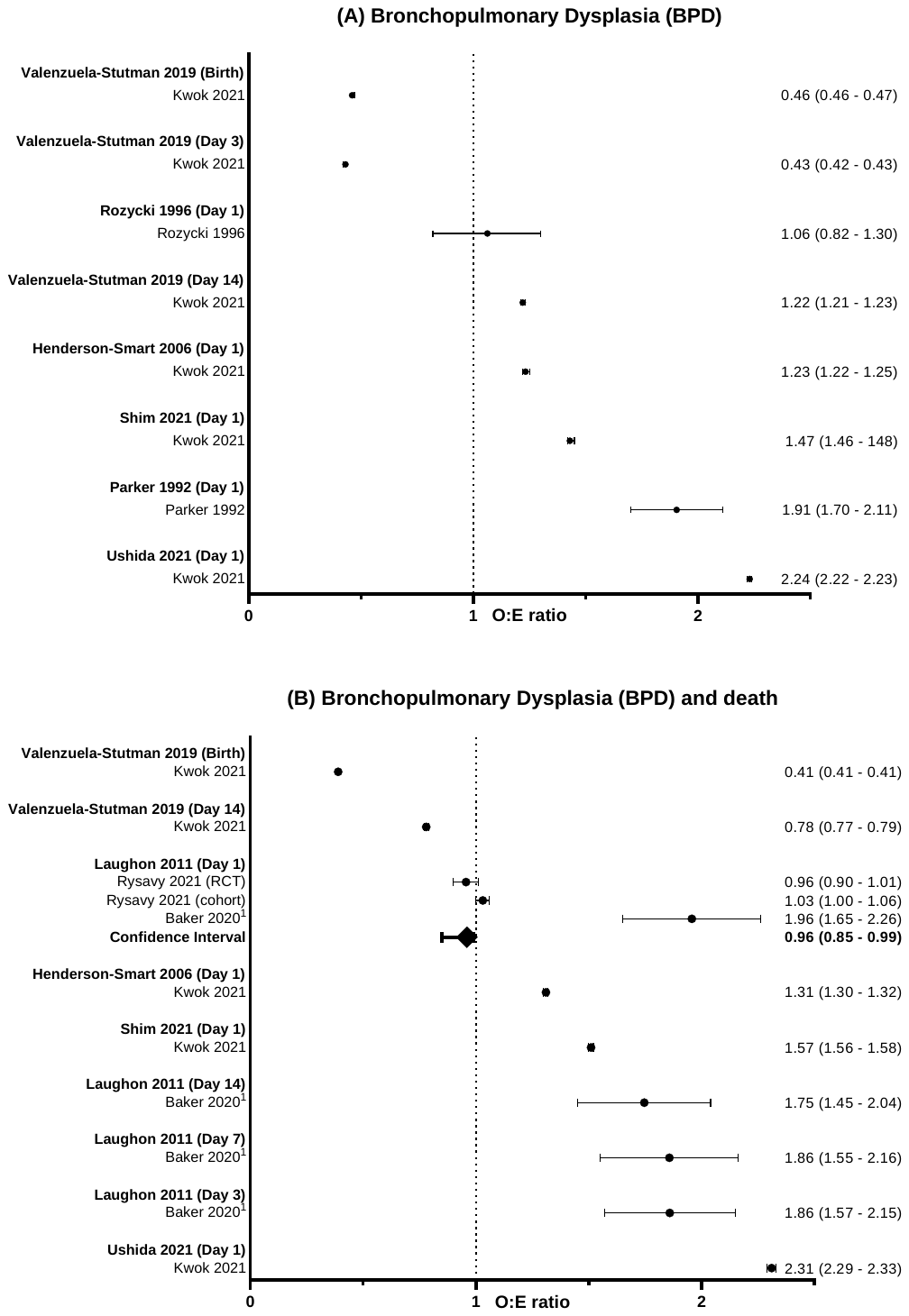

#### Appendix 9: Calibration plots of prediction models with two or more external validation studies.

^1^ Observed and expected risk were obtained from the observed incidence table stratified by estimated risk group.

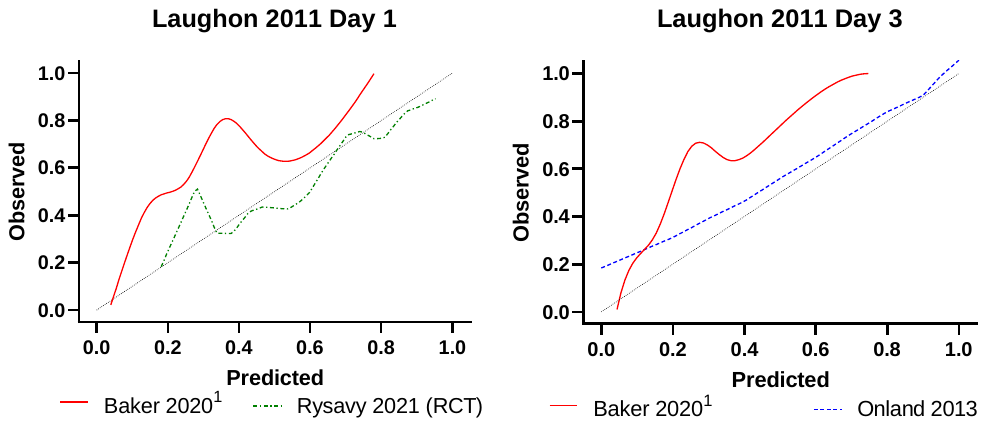

#### Appendix 10: Flow diagram depicting the retrospective patient cohort for the external validation study

Resp = Respiratory. BPD = bronchopulmonary dysplasia

**n = 62,937**

- Born <32 weeks of gestational age
- Born in January 2010 to December 2017
- Admitted to neonatal unit in England & Wales

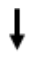

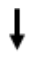

**n = 62,864**

Missing data

- Ethnicity	– 11.8% (n=7,444)
- Apgar at 5 minute	– 10.8% (n=6,796)
- Apgar at 1 minute	– 10.3% (n=6,498)
- Mode of delivery	– 6.1% (n=3,809)
- Antenatal steroids	– 5.6% (n=3,522)
- Admission temperature	– 1.3% (n=803)
- Resp support day 3	– 0.8% (n=474)
- Resp support day 7	– 0.7% (n=453)
- Resp support day 14	– 0.7% (n=425)
- Birthweight z score	– 0.2% (n=150)
- BPD	– 0.6% (n=383)
- Gender	– 0.07% (n=43)
- Outborn/inborn status	– 0.04% (n=24)
- Multiple pregnancy	– 0.02% (n=14)

**Exclusions**

- Birthweight z score above 4 or below -4 (n = 73)

#### Appendix 11: Infant characteristics of the retrospective patient cohort for the external validation study

^1^ Percentage of bronchopulmonary dysplasia was calculated based on babies survived till 36 weeks of corrected gestational age.

| **Clinical characteristics** | **All infants (n = 62,864)** |
| --- | --- |
| **Gestation at birth** (weeks), median (IQR) | 29+3 (27+2 – 30+6) |
| **Birthweight** (g), median (IQR) | 1200 (900 – 1490) |
| **Birthweight z score**, median (IQR) | -0.25 (-0.87 – 0.26) |
| **Small for gestational age** (%) | 9,229 (15) |
| **Sex** (n (%)) |  |
| Male | 34,276 (55) |
| **Ethnicity** (n (%)) |  |
| White  South Asian  Black  Others/Mix | 27,830 (70)  5,138 (13)  3,167 (8)  3,380 (9) |
| **Complete antenatal steroid course** (n (%)) | 42,647 (72) |
| **Apgar at 1 minute** median (IQR) | 6 (4 – 8) |
| **Apgar at 5 minutes** median (IQR) | 9 (7 – 9) |
| **Apgar at 10 minutes** median (IQR) | 9 (8 – 10) |
| **Vaginal delivery** (n (%)) | 24,902 (42) |
| **Received surfactant** (n (%)) | 38,938 (65) |
| **Received invasive ventilation** (n (%)) | 42,385 (67) |
| **Duration of invasive ventilation** (days), median (IQR) | 2 (1 – 7) |
| **Bronchopulmonary dysplasia** (n (%))^1^ | 17,775 (31) |
| **Composite outcome bronchopulmonary dysplasia and death** (n (%)) | 23,095 (37) |
| **Death** (n (%)) | 5,718 (9) |

#### Appendix 12: Decision curve analysis of prediction models using retrospective cohort.

HS = Henderson-Smart. VS = Valenzuela-Stutman

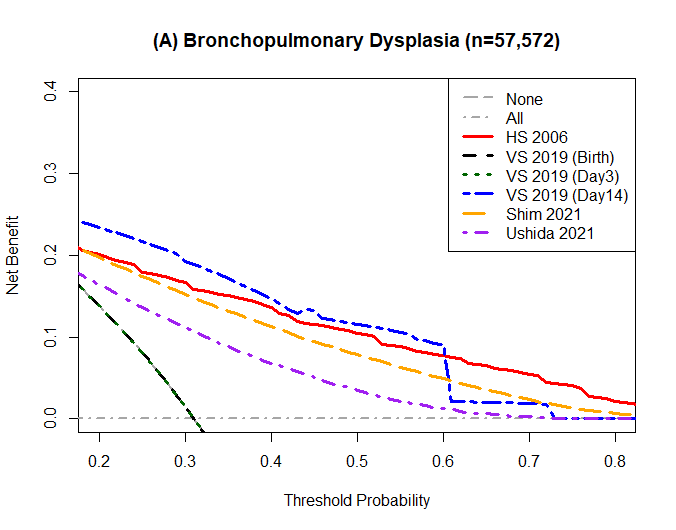

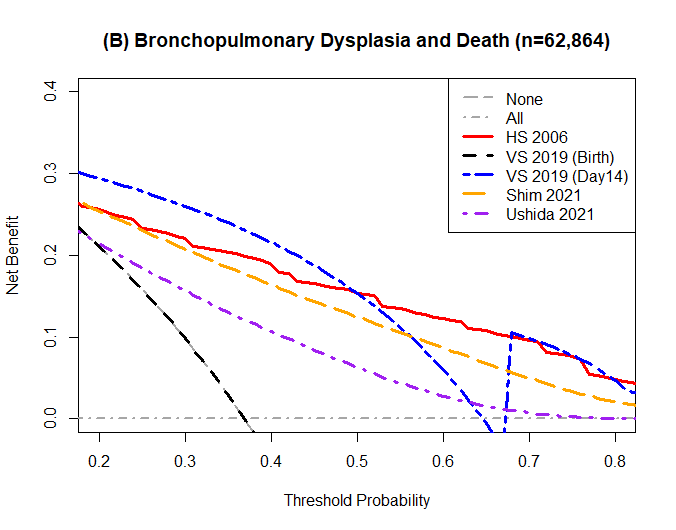

#### Appendix 13: List of participating institutions within the UK Neonatal Collaborative

| **Institution** | **Lead clinician** |
| --- | --- |
| Airedale General Hospital | Dr Matthew Babirecki |
| Arrowe Park Hospital | Dr Anand Kamalanathan |
| Barnet Hospital | Dr Tim Wickham |
| Barnsley District General Hospital | Dr Kavi Aucharaz |
| Basildon Hospital | Dr Aashish Gupta |
| Basingstoke & North Hampshire Hospital | Dr Nicola Paul |
| Bassetlaw District General Hospital | Dr L M Wong |
| Bedford Hospital | Dr Anita Mittal |
| Birmingham City Hospital | Dr Lindsay Halpern |
| Birmingham Heartlands Hospital | Dr Pinki Surana |
| Birmingham Women's Hospital | Dr Matt Nash |
| Bradford Royal Infirmary | Dr Sam Wallis |
| Broomfield Hospital, Chelmsford | Dr Ahmed Hassan |
| Calderdale Royal Hospital | Dr Karin Schwarz |
| Chelsea & Westminster Hospital | Dr Shu-Ling Chuang |
| Chesterfield & North Derbyshire Royal Hospital | Dr Aiwyne Foo |
| Colchester General Hospital | Dr Jo Anderson |
| Conquest Hospital | Dr Graham Whincup |
| Countess of Chester Hospital | Dr Stephen Brearey |
| Croydon University Hospital | Dr Morris |
| Croydon University Hospital | Dr Srirambhatla |
| Cumberland Infirmary | Dr Yee Aung |
| Darent Valley Hospital | Dr Abdul Hasib |
| Darlington Memorial Hospital | Dr Mehdi Garbash |
| Derriford Hospital | Dr Alex Allwood |
| Diana Princess of Wales Hospital | Dr Pauline Adiotomre |
| Doncaster Royal Infirmary | Dr Nigel Brooke |
| Dorset County Hospital | Dr Abby Deketelaere |
| East Surrey Hospital | Dr Abdul Khader |
| Epsom General Hospital | Dr Sonia Spathis |
| Frimley Park Hospital | Dr Sanghavi Rekha |
| Furness General Hospital | Dr Anas Olabi |
| George Eliot Hospital | Dr Mukta Jain |
| Glan Clwyd Hospital | Dr Ian Barnard |
| Glangwili General Hospital | Dr Prem Pitchaikani |
| Gloucester Royal Hospital | Dr Jennifer Holman |
| Good Hope Hospital | Dr Pinki Surana |
| Great Western Hospital | Dr Stanley Zengeya |
| Guy's & St Thomas' Hospital | Dr Geraint Lee |
| Harrogate District Hospital | Dr Sobia Balal |
| Hereford County Hospital | Dr Cath Seagrave |
| Hillingdon Hospital | Dr Tristan Bate |
| Hinchingbrooke Hospital | Dr Hilary Dixon |
| Homerton Hospital | Dr Narendra Aladangady |
| Hull Royal infirmary | Dr Hassan Gaili |
| Ipswich Hospital | Dr Matthew James |
| James Cook University Hospital | Dr M Lal |
| James Paget Hospital | Dr Ambadkar |
| Kettering General Hospital | Dr Poornima Pandey |
| Kings College Hospital | Dr Ravindra Bhat |
| King's Mill Hospital | Dr Simon Rhodes |
| Kingston Hospital | Dr Jonathan Filkin |
| Lancashire Women and Newborn Centre | Dr Savi Sivashankar |
| Leeds Neonatal Service | Dr Lawrence Miall |
| Leicester General Hospital | Dr Jonathan Cusack |
| Leicester Royal Infirmary | Dr Venkatesh Kairamkonda |
| Leighton Hospital | Dr Michael Grosdenier |
| Lincoln County Hospital | Dr Ajay Reddy |
| Lister Hospital | Dr J Kefas |
| Liverpool Women's Hospital | Dr Christopher Dewhurst |
| Luton & Dunstable Hospital | Dr Jennifer Birch |
| Macclesfield District General Hospital | Dr Gail Whitehead |
| Manor Hospital | Dr Ashok Karupaiah |
| Medway Maritime Hospital | Dr Ghada Ramadan |
| Milton Keynes General Hospital | Dr I Misra |
| Musgrove Park Hospital | Dr Chris Knight |
| New Cross Hospital | Dr Matt Nash |
| Newham General Hospital | Dr Imdad Ali |
| Nobles Hospital | Dr Prakash Thiagarajan |
| Norfolk & Norwich University Hospital | Dr Muthukumar |
| North Devon District Hospital | Dr Michael Selter |
| North Manchester General Hospital | Dr Ajit Mahaveer |
| North Middlesex University Hospital | Dr Neeraj Jain |
| Northampton General Hospital | Dr Subodh Gupta |
| Northumbria Specialist Emergency Care Hospital | Jess Reynolds |
| Northwick Park Hospital | Dr Richard Nicholl |
| Nottingham City Hospital | Dr Steven Wardle |
| Nottingham University Hospital (QMC) | Dr Steven Wardle |
| Ormskirk District General Hospital | Dr Andreea Bontea |
| Oxford University Hospitals, John Radcliffe Hospital | Dr Eleri Adams |
| Peterborough City Hospital | Dr Katharine McDevitt |
| Pilgrim Hospital | Dr Ajay Reddy |
| Pinderfields General Hospital (Pontefract General Infirmary) | Dr David Gibson |
| Poole General Hospital | Prof Minesh Khashu |
| Prince Charles Hospital | Dr Iyad Al-Muzaffar |
| Princess Alexandra Hospital | Dr Chinnappa Reddy |
| Princess Anne Hospital | Dr Mark Johnson |
| Princess of Wales Hospital | Dr Kate Creese |
| Princess Royal Hospital | Dr P Amess |
| Princess Royal Hospital (previously Royal Shrewsbury Hospital) | Dr Deshpande |
| Princess Royal University Hospital | Dr Elizabeth Sleight |
| Queen Alexandra Hospital | Dr Charlotte Groves |
| Queen Charlotte's Hospital | Dr Lidia Tyszcuzk |
| Queen Elizabeth Hospital, Gateshead | Dr Anne Dale |
| Queen Elizabeth Hospital, King's Lynn | Dr Glynis Rewitzky |
| Queen Elizabeth Hospital, Woolwich - see notes | Dr Olutoyin Banjoko |
| Queen Elizabeth the Queen Mother Hospital | Dr Bushra Abdul-Malik |
| Queen's Hospital, Burton on Trent | Dr Dominic Muogbo |
| Queen's Hospital, Romford | Dr Khalid Mannan |
| Queen's Hospital, Romford 2 | Dr Khalid Mannan |
| Rosie Maternity Hospital, Addenbrookes | Dr Angela D'Amore |
| Rotherham District General Hospital | Dr Soma Sengupta |
| Royal Albert Edward Infirmary | Dr Christos Zipitis |
| Royal Berkshire Hospital | Dr Peter De Halpert |
| Royal Bolton Hospital | Dr Paul Settle |
| Royal Cornwall Hospital | Dr Paul Munyard |
| Royal Derby Hospital | Dr John McIntyre |
| Royal Devon & Exeter Hospital | Dr Chrissie Oliver |
| Royal Gwent Hospital | Dr Sunil Reddy |
| Royal Hampshire County Hospital | Dr Lucinda Winckworth |
| Royal Lancaster Infirmary | Dr Joanne Fedee |
| Royal Oldham Hospital | Dr Natasha Maddock |
| Royal Preston Hospital | Dr Richa Gupta |
| Royal Stoke University Hospital | Dr Jyoti Kapur |
| Royal Surrey County Hospital | Dr Ben Obi |
| Royal Sussex County Hospital | Dr P Amess |
| Royal United Hospital | Dr Stephen Jones |
| Royal Victoria Infirmary | Dr Naveen Athiraman |
| Russells Hall Hospital | Dr Chandan Gupta |
| Salisbury District Hospital | Dr Jim Baird |
| Scarborough General Hospital | Dr Kirsten Mack |
| Scunthorpe General Hospital | Dr Pauline Adiotomre |
| Singleton Hospital | Dr Arun Ramachandran |
| Southend Hospital | Dr Vineet Gupta |
| Southmead Hospital | Dr Faith Emery |
| St George's Hospital | Dr Charlotte Huddy |
| St Helier Hospital | Dr Ralf Hartung |
| St Mary's Hospital, IOW | Dr Akinsola Ogundiya |
| St Mary's Hospital, London | Dr Lidia Tyszcuzk |
| St Mary's Hospital, Manchester | Dr Ngozi Edi-Osagie |
| St Michael's Hospital | Dr Pamela Cairns |
| St Peter's Hospital | Dr Peter Martin |
| St Richard's Hospital | Dr Victoria Sharp |
| Stepping Hill Hospital | Dr Carrie Heal |
| Stoke Mandeville Hospital | Dr Sanjay Salgia |
| Sunderland Royal Hospital | Dr Majd Abu-Harb |
| Tameside General Hospital | Dr Jacqeline Birch |
| The Grange University Hospital | Dr Sunil Reddy |
| The Jessop Wing, Sheffield | Dr Porus Bastani |
| The Royal Free Hospital | Dr Marice Theron |
| The Royal London Hospital - Constance Green | Dr Vadivelam Murthy |
| Torbay Hospital | Dr Siba Paul |
| Tunbridge Wells Hospital | Dr Hamudi Kisat |
| University College Hospital | Dr Giles Kendall |
| University Hospital Coventry | Dr Puneet Nath |
| University Hospital Lewisham | Dr Ozioma Obi |
| University Hospital of North Durham | Dr Mehdi Garbash |
| University Hospital of North Tees | Dr Hari Kumar |
| University Hospital of Wales | Dr Nitin Goel |
| Victoria Hospital, Blackpool | Dr Chris Rawlingson |
| Warrington Hospital | Dr Delyth Webb |
| Warwick Hospital | Dr Bird |
| Watford General Hospital | Dr Sankara Narayanan |
| West Cumberland Hospital | Dr Yee Aung |
| West Middlesex University Hospital | Dr Eleanor Hulse |
| West Suffolk Hospital | Dr Ian Evans |
| Wexham Park Hospital | Dr Sanjay Jaisal |
| Whipps Cross University Hospital | Dr Caroline Sullivan |
| Whiston Hospital | Dr Ros Garr |
| Whittington Hospital | Dr Wynne Leith |
| William Harvey Hospital | Dr Vimal Vasu |
| Withybush Hospital | Dr Vishwa Narayan |
| Worcestershire Royal Hospital | Dr Liza Harry |
| Worthing Hospital | Dr Katia Vamvakiti |
| Wrexham Maelor Hospital | Dr Brendan Harrington |
| Wythenshawe Hospital | Dr Ngozi Edi-Osagie |
| Yeovil District Hospital | Dr Megan Eaton |
| York District Hospital | Dr Sundeep Sandhu |
| Ysbyty Gwynedd | Dr Mike Cronin |

List was accessed from <https://www.imperial.ac.uk/neonatal-data-analysis-unit/neonatal-data-analysis-unit/list-of-national-neonatal-units/> on 06/01/2022.
